# The Birth of Influenza Immunity: High-Resolution Antibody Dynamics Driven by Maternal Antibody Waning, Vaccinations, and Infections during the first Two Years of Life

**DOI:** 10.64898/2026.05.04.26352413

**Authors:** Soo Min Lee, Allison R. Burrell, Sara Spranger, Shannon C. Conrey, Brendon White, Ardythe L. Morrow, Daniel C. Payne, Mary Allen Staat, Tal Einav

## Abstract

Although infants are often described as immunological clean slates, the magnitude and durability of their first immune responses, and their effects on subsequent exposures, remains unclear. We conducted a longitudinal study of *n*=245 children across their first two years of life, mapping influenza antibody dynamics following vaccinations and infections. Maternal antibodies efficiently transferred to infants but exhibited different kinetics, with influenza B antibodies persisting longer than influenza A (half-life≈75 vs 50 days). Among infants whose maternal antibodies did not fully decay by the time of their 1st vaccination, 83-94% had no influenza A response while 59-72% had no influenza B response. To account for such maternal interference in infant vaccine responses, we formulated a personalized clinical algorithm that uses a single maternal blood draw and antibody half-lives to predict when maternal antibodies will reach undetectable levels. In addition, antibody responses varied across vaccine strains, with most infants exhibiting no response to H1N1 or H3N2 during 1st vaccination but eliciting a response after 2nd vaccination. Consequently, infants that turned 6 months old and were vaccinated in February-May ‒ after the peak in influenza cases ‒ showed enhanced antibody titers the following season, suggesting that ∼33% of infants in this category may benefit from late-season vaccination. Across the 60 infections captured, influenza A infections elicited strong, subtype-specific responses while influenza B led to weaker but more cross-reactive responses. Irrespective of prior infections, a strong 1st vaccine response was a predictor of a strong 2nd vaccine response, suggesting that the 1st vaccine response influences subsequent influenza immunity.

## Introduction

Influenza vaccination remains the most effective strategy to prevent influenza morbidity and mortality, yet immune responses vary widely across the population. Each influenza season, roughly 67% of adult vaccinees mount little-to-no antibody response as measured by hemagglutination inhibition (HAI), while only 16% develop durable protection lasting to the next influenza season.^1–5^ Longitudinal studies found that most individuals alternate between strong and weak responses across seasons,^6^ but the mechanisms driving this heterogeneity remain poorly understood.

Infants offer a rare opportunity to examine the influenza antibody response without the confounding effects of prior infections or vaccinations. Early influenza immunity is built upon maternal antibodies transferred during gestation that wane over the first months of life, and prior work has examined facets of this infant antibody response. Cord blood titers were found to be 1.1-2.3x larger than maternal titers during delivery.^7–10^ These maternally-derived antibodies then wane with a half-life of 28-95 days,^9,11^ with most infants reaching the limit-of-detection for H1N1 and influenza B (HAI geometric mean titer, GMT≈5) and very low H3N2 titers (GMT≈5-10) at 6 months,^12–14^ the age at which they can receive their first influenza vaccination.

Other work examined the immunogenicity of infant vaccine responses and revealed dramatic variability in antibody titers. A four-year vaccine study from 1985-1988, when the H3N2 vaccine strain changed each year, reported that 6-18 month old infants had pre→post-vaccination titers of 5→11 in 1985, 5→380 in 1986, 5→15 in 1987, and 5→51 in 1988 (fold-change range: 2.2-76x).^12^ Vaccine studies in 2004-2005, where the H3N2 vaccine strain was again updated, reported pre→post-vaccination GMTs in infants of 13→21 in 2004 and 5→40 in 2005 (fold-change range: 1.6-8x).^13^ Both studies found substantially lower influenza B titers. Thus, despite having little-to-no prior exposures, “clean slate” infant immunity responded very differently to different antigens.

One limitation of prior studies is that they have not systematically examined all of these effects in the same infants. Here, we present influenza serological responses from the PREVAIL (Pediatric Respiratory & Enteric Virus Acquisition and Immunogenesis Longitudinal) cohort, a prospective observational maternal-birth cohort that enrolled *n*=245 maternal-child pairs in the Cincinnati, Ohio metropolitan area on a rolling basis from 2017 to 2018 and assessed their antibody responses over their first two years of life. Whereas most vaccine studies align sample collection to the date of vaccination, PREVAIL sample collection was based upon each child’s age, providing a much broader sampling of pre- and post-vaccination as well as pre- and post-infection time points. As a result, this work maps out the dynamics of infant vaccine responses with high-resolution that has thus far only been possible in adults by integrating multiple vaccine studies.^2–5,15^

Infections in PREVAIL children were monitored through weekly nasal swabs with influenza RT-PCR and/or rapid influenza diagnostic tests performed during medical visits, providing a rare opportunity to not only examine antibody titers following infection but also quantify the “clean” vaccination response without prior infections. Stratifying participants by prior exposure history, this work investigates whether the antibody ceiling effect extends to infants and children^16,17^ or if influenza infections prime subsequent vaccination responses as seen in infants and adults.^3,18,19^ Moreover, we examine questions that are unique to this stage of life such as whether infants that turn six months old in February-May (late in the northern hemisphere’s influenza season when there are typically fewer influenza infections) benefit from vaccination during this time.

Collectively, this work describes the serological profiles needed to better understand and enhance antibody responses in infants and children.

## Results

### 1) The PREVAIL cohort combined standardized serology with in-depth monitoring of influenza vaccinations and infections during the first two years of life

From the *n*=245 children in the PREVAIL cohort (**Table 1**), serum was collected at 0 (cord), 1.5, 6, 12, 18, and 24 months after birth, and antibody titers were measured using hemagglutination inhibition (HAI) against the contemporary influenza vaccine strains from that season (**Fig S1**). Each child’s full vaccination history and their mother’s vaccination history during the 18 months prior to delivery were captured.

**Table 1.** Demographic information for the PREVAIL cohort (*n*=245).

| Variable | PREVAIL Statistic |
| --- | --- |
| Gestational Age (weeks; mean ± SD) | 38.8 ± 1.2 |
| Maternal Age at Birth (years; mean ± SD) | 29.2 ± 5.3 |
| Maternal Race (White / Black / Other) | 51.4% / 43.7% / 4.9% |
| Maternal Ethnicity (Non-Hispanic / Hispanic) | 97.6% / 2.4% |
| Duration of Breastfeeding* (days; mean ± SD) | 179 ± 199 |
| Infant Sex (Female / Male) | 51.8% / 48.2% |
| Number of Infant Influenza Vaccines from 0-2 y.o. (0 / 1 / ≥2) | 29.8% / 10.6% / 59.6% |
| Number of Infant Influenza Infections from 0-2 y.o.** (0 / 1 / ≥2) | 78.0% / 19.2% / 2.9% |

\* Counts exclusive breastfeeding as well as partial breastfeeding supplemented by formula.
\*\* Determined through nasal swabs and influenza tests from medical visits.

Children receiving their first influenza vaccine are recommended to get two doses of the vaccine at least one month apart, and if they do not, they are recommended to get two doses the following season. Because sera were collected in 6 month intervals, which typically includes the administration of both doses, our study design cannot examine the effects of each vaccine dose. Instead, in this work we define the antibody response during the *1st vaccination season* (henceforth referred to as the 1st vaccine response) as the response to all doses (either one or two) received in the influenza season containing their first vaccine dose (the influenza season spans July 1 until the following June 30, **Methods**). Similarly, we define the antibody response during the *2nd vaccination season* (hereafter referred to as the 2nd vaccine response) as all doses given in the closest subsequent influenza season. Thus, antibody titers following the 1st vaccine response include the 114 infants that received two doses as well as the 39 infants that received a single dose.

Influenza infections were monitored through nasal swabs that were collected weekly, regardless of symptoms, in order to capture both asymptomatic and symptomatic infections; swabs were sequenced to determine the subtype or lineage of the infecting strain (H1N1, H3N2, B Victoria, or B Yamagata). Infections were additionally determined through electronic health records from all medical visits where the children had a fever or cough, and rapid influenza diagnostic tests during such visits determined the type of the infecting strain (influenza A or influenza B).

Using this deeply phenotyped cohort, we quantified the antibody response elicited by influenza vaccination in the first years of life as well as the impact of infections. In the following sections, we proceed chronologically from birth through the 1st and 2nd vaccine responses, first analyzing “clean” vaccine response by excluding titers from infected children (in the homologous infection subtype/lineage only, **Methods**), and in the final section examining the impact of infections. Overall adherence was high across the cohort, with 79% of intended serum samples collected and 48% of intended weekly nasal swab collections completed over the follow-up period, and the following analyses use all available data.

### 2) Maternal antibodies against all influenza subtypes were passed to infants, with the highest titers seen in infants whose mother was vaccinated during pregnancy

Serum was collected from pregnant mothers in the third trimester (≥34 weeks gestation), resulting in 221 cases where maternal antibody titers could be compared against infant cord blood titers (**Fig 1A**). These paired titers showed a near perfect match across the entire cohort and all vaccine strains, with root-mean-squared error (RMSE) of 1.8x, 1.9x, 1.8x, 1.8x for H1N1, H3N2, B Victoria, and B Yamagata, respectively, comparable to the ≈2x intrinsic noise of the HAI assay (**Fig 1B**). These data indicate that maternal antibodies are efficiently transferred to infants, resulting in cord GMTs of ≈120 for H1N1 and H3N2, 70 for B Victoria, and 160 for B Yamagata (**Fig S2**).

**Figure 1.**
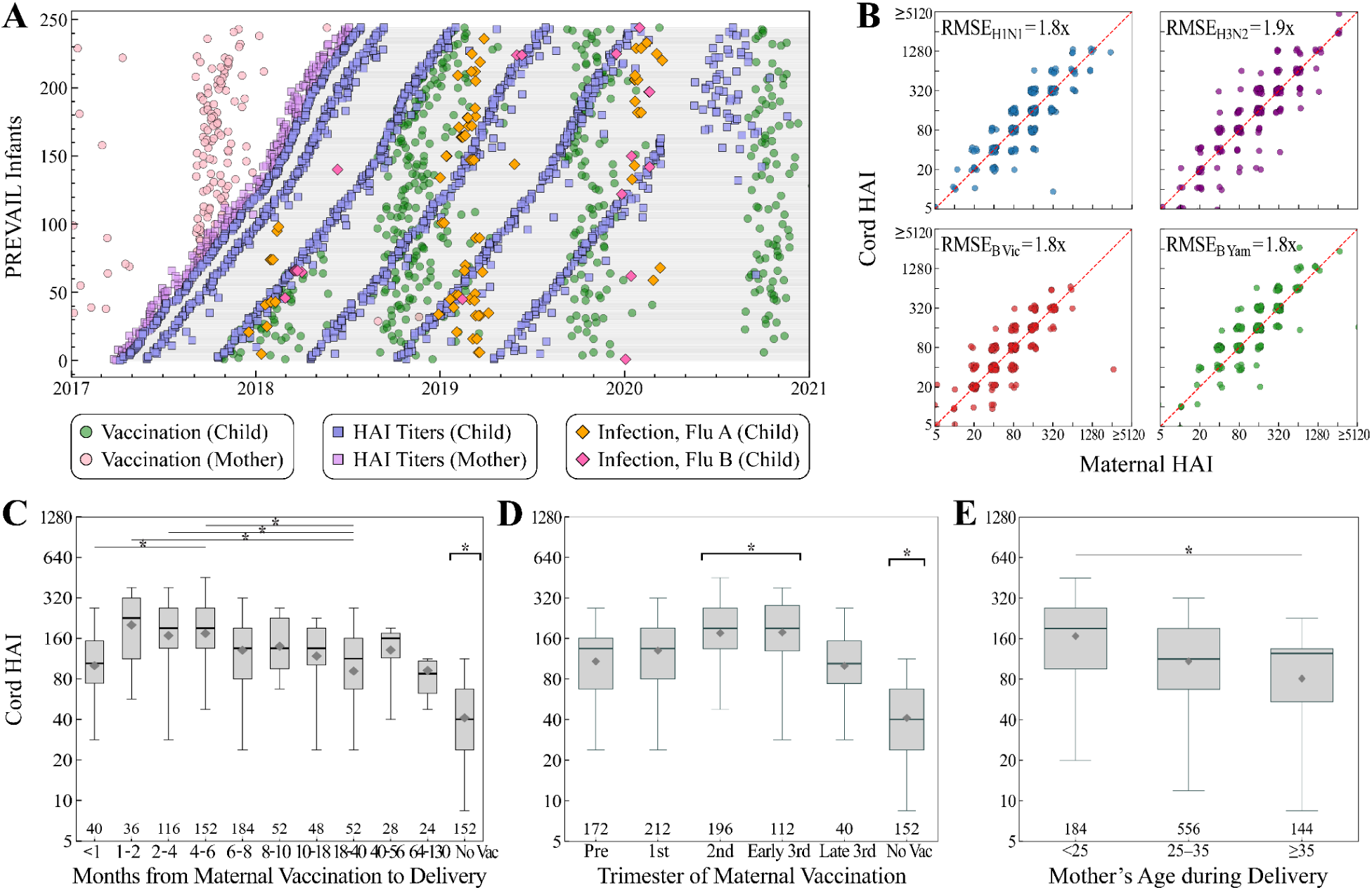
Dependence of maternal and infant cord blood titers on maternal vaccination. (A) Timeline of the PREVAIL cohort, with subjects ordered chronologically by date of birth. Influenza vaccinations (circles), HAI titers (squares), and infant infections (diamonds) were extensively tracked across the years shown. (B) Maternal vs infant cord blood HAI titers during delivery for each vaccine strain. Dependence of each infant’s cord blood GMT from all vaccine strains on (C) time from the most recent maternal influenza vaccine to delivery [1 month=30 days], (D) time from start of pregnancy to maternal vaccination, and (E) maternal age during delivery. In Panels C-D, some mothers had no vaccine in the prior 18 months (No Vac). In Panel D, maternal vaccination could fall within 18 months of delivery but pre-conception (Pre). Vaccination during the 3rd trimester was split into its first two months (Early) on its final month (Late). All box plots show the interquartile range, horizontal lines the medians, and black diamonds the GMTs. Whiskers extend to 1.5 times the interquartile range. The number of measurements=(# of mothers)×(# of vaccine strains) are shown for each box plot. Bracket indicates significance against all other groups. *: *p* ≤ 0.05 (Welch’s t-test on the log-transformed values).

Influenza vaccination before delivery should boost both the mother’s antibody titers and the infant’s titers during their first months of life, thereby enhancing their protection.^20^ Indeed, among these mother-infant pairs, 83% of mothers had a documented history of influenza vaccination prior to delivery. Vaccination during the 2nd or 3rd trimester, but at least 30 days prior to delivery, led to ∼2x larger maternal (and cord blood) titers compared to vaccinating at any other time (**Fig 1C,D**). In addition, mothers not vaccinated in the past 18 months had significantly decreased titers with GMTs ∼2x lower than any vaccinated group (*p*≤0.05, Welch’s t-test) (**Fig 1C,D**). This trend remained consistent across individual vaccine strains and maternal age groups (<25, 25–35, and ≥35 years), confirming that these timing-dependent differences were not confounded by strain or age (**Fig S3**).

Independent of vaccination timing, maternal age at delivery also significantly affected titers, with mothers <25 y.o. exhibiting 2x larger titers than mothers >35 y.o. (*p*≤0.05, Welch’s t-test), while mothers 25-35 had intermediate titers (**Fig 1E**). Notably, this same transition of 2x lower titers between ages 25-35 mirrors the age dependence of males and females from dozens of influenza vaccine studies and influenza seasons,^17^ and hence is not restricted to pregnant mothers.

### 3) Maternal antibodies waned with predictable half-lives, with influenza B antibodies decaying slower than influenza A

After birth, maternal antibodies decayed at a constant rate for each vaccine component, consistent with exponential decay (**Fig 2A**). Influenza B antibodies decayed far slower than influenza A antibodies, with half-lives of 50 days for H1N1, 49 days for H3N2, 80 days for B Victoria, and 71 days for B Yamagata, as inferred through linear regression (**Methods**).

**Figure 2.**
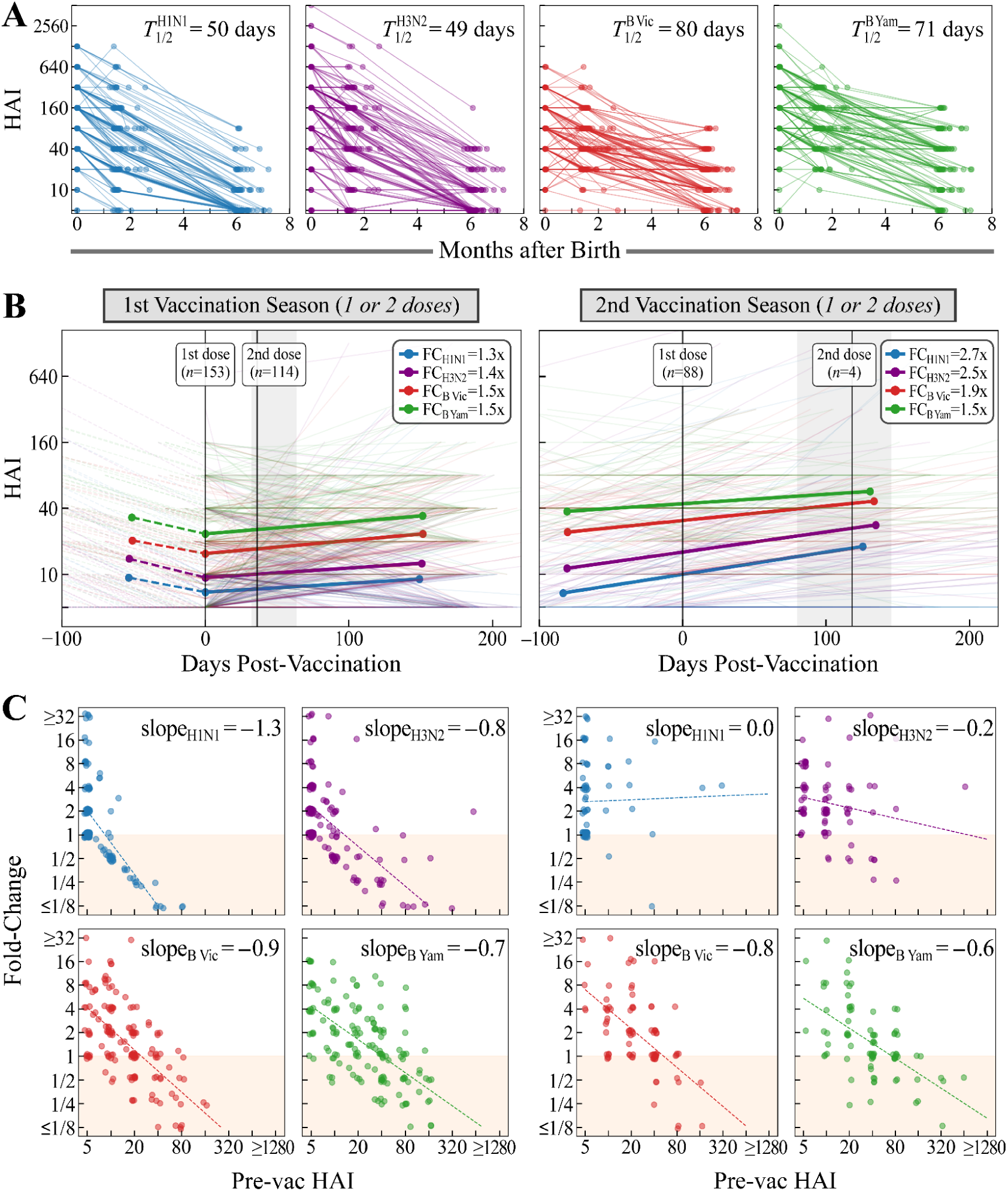
Responses to 1st and 2nd vaccination in the absence of infection. (A) Waning of infant antibodies during the first months of life. The half-life (*T*_1/2_) for each vaccine component is inferred through linear regression. (B) Antibody titers from each infant’s 1st and 2nd vaccination seasons (each considering all doses received during an influenza season, **Methods**). For the 1st vaccine, titers were imputed (dashed lines) from the last measured titers to the day of vaccination using the half-life of maternal antibodies, and fold-change was measured relative to these titers. The mean and 50% quartile range for each vaccine dose, and the number of infants receiving a second dose, are shown by a vertical line and shading. Legends show fold-change (FC=post-vaccination titer/pre-vaccination titer) for each vaccine strain. (C) The antibody ceiling effect for each vaccine strain quantifying the slope between pre-vaccination titer and fold-change. Points are jittered for clarity, and the shaded area denotes no vaccine response (FC≤1).

By six months of age, when infants can receive their first influenza vaccine, B Victoria and B Yamagata titers decayed to ∼1/5 of cord blood titers on average, with <10% of infants reaching the limit of detection (HAI=5). In contrast, titers for H1N1 and H3N2 declined to ∼1/17 of cord blood levels on average, with 47% of infants reaching the detection limit. For example, all infants with cord blood HAI ≤80 for H1N1 reached HAI of 5 by six months.

Interestingly, a subset of 35/67 infants that were not vaccinated during their first year of life had titers that remained above the limit of detection for both influenza B strains (HAIs=10-40, **Fig S4B**). Such a plateau could artificially increase the influenza B titer half-life at the later time points, yet refitting half-lives using only the 0 and 1.5 month titers led to comparable half-lives with influenza A decaying significantly faster than influenza B. Altogether, this demonstrated that all maternally-derived influenza titers decayed during the first months of life, yet influenza B titers were significantly more durable.

### 4) The 1st influenza vaccine elicited substantially weaker antibody titers than the 2nd vaccine

One barrier to quantifying each child’s 1st vaccine response in a longitudinal study is the variability in pre-vaccination collection dates. On average, serum was collected 50 days prior to vaccination (range: 405 days prior to 13 days after, **Fig S6A**), and directly using these measured titers would not account for maternal antibody decay. Instead, the antibody half-lives found above were used to extend the pre-vaccination measurements forward in time to impute each child’s titers on the day of vaccination (**Methods**). In effect, this imputation transformed the longitudinal PREVAIL study into a classic vaccine study where HAI titers are measured right before vaccination.

Across all children, *n*=153 had sera collected before and after their 1st vaccination (**Fig S5**, **Fig S6A**). Using their imputed pre-vaccination titers and the earliest post-vaccination measurement, this cohort exhibited a modest but statistically significant fold-change of 1.3x fold-change for H1N1, 1.4x for H3N2 and 1.5x for both influenza B viruses (*p*≤0.05, paired t-test) (**Fig 2B**, left). As a point of comparison, all fold-changes were far smaller than the ∼3x fold-change seen in adults for each vaccine strain in most influenza seasons.^17^

Due to the faster decay of maternal influenza A antibodies, influenza B titers were higher than influenza A titers pre-1st-vaccination. This hierarchy was maintained post-1st-vaccination, where influenza B GMTs of 23-34 were significantly higher than the influenza A GMTs of 9-13 (*p*≤0.05, Welch’s t-test). Although current guidelines suggest that the 1st influenza vaccine should be administered as two full doses 1 month apart, neither the receipt nor the timing of this second dose was enforced during the PREVAIL study, and hence the impact of the second dose could be evaluated retrospectively. The 1.4x fold-change observed in the *n*=114 infants who received a second dose was not significantly different from the 1.5x fold-change seen in the *n*=39 infants who received only a single dose (*p*=0.77, Welch’s t-test), although this sample size was too small to test the impact of the time between both doses.

Interestingly, these 1st vaccine responses showed a far stronger but more variable antibody ceiling effect than seen in adults, as quantified by the slope of fold-change vs pre-vaccination HAI. All four vaccine strains showed pronounced diagonal bands (**Fig 2C**, left), with the bottom-left band denoting children with post-vaccination titer=5, the next band post-vaccination titer=10, and so on, with the influenza B viruses displaying more bands and thus more heterogeneity than influenza A viruses. Linear regression of the H1N1, H3N2, and B Victoria responses all showed a hard ceiling for each band (*i.e.*, 2x larger pre-vac titer results in 2x smaller fold-change), while B Yamagata showed a weaker effect (4x larger pre-vaccination titer results in 2x smaller fold-change). In comparison, the antibody ceiling effect appears as a single band with a far milder slope in adults, where a 10x increase in pre-vaccination titers was required to decrease fold-change by 2x.^6^ To demonstrate that this strong antibody ceiling effect was not an artifact of the continuous imputations nor maternal antibody waning, data was refit without these features, yet the same multi-banded hard ceiling was found each time (**Fig S7**).

We next assessed the 2nd vaccine response in the *n*=88 infants with sera collected pre- and post-vaccination (**Fig S6B**). Titers for H3N2 and both B viruses were nearly identical between the post-1st-vaccine and pre-2nd-vaccine measurements, with only H1N1 showing a small decline from GMT of 9→7 (**Fig 2B**). Compared to the 1st vaccination, the antibody response to the 2nd vaccination was far larger with a fold-change of 2.7x for H1N1 and 2.5x for H3N2 (*p*≤0.05, paired t-test) (**Fig 2B**, right). Although the B viruses had fold-change≈1.7x, comparable to the 1st vaccine, the post-vaccination GMTs of the B viruses continued to be larger than the influenza A GMTs (*p*≤0.05, Welch’s t-test). While *n*=4 infants received a two-dose regimen for the 2nd vaccine, their responses did not noticeably differ from those who received only a single dose (*p*=0.43, Welch’s t-test).

The antibody ceiling effect was slightly weaker for influenza B and far weaker for influenza A during 2nd vaccination. More precisely, a 2x decrease in fold-change resulted from 3-4x increased pre-vaccination titers for the B viruses or 4-5x increased titers against H3N2 (**Fig 2C**, right). H1N1 exhibited a zero slope (an anti-ceiling), although 61 out of 75 infants had pre-vaccination HAI=5, making this slope highly uncertain.

During both the 1st and 2nd vaccination, there was dramatic heterogeneity in the responses of individual infants, with 66% of infants exhibiting ≥4x fold-change against at least one strain in either vaccine. Of the infants with ≥4x fold-change against one vaccine strain, their geomean fold-change to the other vaccine strains was always <4x during the 1st vaccination (**Table 2, Fig S8B**). This suggests that the antibody response to each strain was essentially independent, and such biases could arise each infant’s HLA type.^21^

**Table 2.** Geometric mean fold-change against subtypes for strong responders for (A) 1st and (B) 2nd vaccination. HAI fold-change following (A) 1st and (B) 2nd vaccination. For all children with fold-change ≥4x against one subtype- or lineage-of-interest [left column], the geometric mean of their fold-change against each vaccine strain [middle columns; subtype-/lineage-of-interest shaded grey] or against the three other vaccine strains [right column; excluding the subtype-/lineage-of-interest]. Cases where fold-change ≥4x outside the subtype-/lineage-of-interest are highlighted in green.

| $\geq 4x$ Fold-Change against | Fold-Change H1N1 | Fold-Change H3N2 | Fold-Change B Vic | Fold-Change B Yam | Fold-Change against All Other Subtypes |
| --- | --- | --- | --- | --- | --- |
| H1N1<br>[n=29] | 8.9x | 2.2x | 3.1x | 3.1x | 2.8x |
| H3N2<br>[n=30] | 2.4x | 8.8x | 3.7x | 3.5x | 3.2x |
| B Victoria<br>[n=37] | 2.2x | 3.2x | 7.0x | 3.1x | 2.8x |
| B Yamagata<br>[n=40] | 2.6x | 3.2x | 3.9x | 6.4x | 3.2x |

**B**
| $\geq 4x$ Fold-Change against | Fold-Change H1N1 | Fold-Change H3N2 | Fold-Change B Vic | Fold-Change B Yam | Fold-Change against All Other Subtypes |
| --- | --- | --- | --- | --- | --- |
| H1N1<br>[n=31] | 9.0x | 3.7x | 3.0x | 2.8x | 3.2x |
| H3N2<br>[n=30] | 4.6x | 7.0x | 2.9x | 2.4x | 3.2x |
| B Victoria<br>[n=27] | 3.8x | 4.8x | 6.9x | 3.4x | 3.9x |
| B Yamagata<br>[n=20] | 6.2x | 4.7x | 5.0x | 6.7x | 5.3x |

The 2nd vaccination elicited an asymmetrical response to influenza A and B. Infants with ≥4x fold-change against influenza A showed weaker fold-change<4x against influenza B, whereas infants with ≥4x fold-change against influenza B also showed stronger fold-change (range: 3.8-6.2x) against influenza A (**Table 2**). Strong B Yamagata responses elicited ≥4x fold-change against every other vaccine strain, while strong responses against B Victoria and H3N2 led to strong responses against a subset of other vaccine strains (**Table 2**).

Only 32% of children with ≥4x fold-change against any strain during their 1st vaccination were likely to have ≥4x fold-change to that same strain during their 2nd vaccination (**Fig S8C**). In contrast, 81% of children with HAI titer≥40 post-1st-vaccination maintained a titer≥40 post-2nd-vaccination (**Fig S8D-F**), comparable to the 82% of adults that maintained this titer level across seasons.^6^ Taken together, infants generally mounted a weak 1st vaccine response and stronger 2nd vaccine response, with the few exceptionally strong responses typically subtype- or lineage-specific.

### 5) Should infants turning 6 months old at the end of the influenza season get vaccinated that season?

In the US, infants can receive their first influenza vaccine after they turn 6 months old,^22^ and influenza case counts typically peak from December through February. Thus, infants that turn 6 months old after February often forgo vaccination until the following season. Yet the preceding analysis suggested that receiving the 1st vaccination, even if in March-June (before these vaccines expired on June 30), should enhance the influenza response in the subsequent season. Indeed, the HAI trajectories of infants that did or did not receive a late-season vaccination showed that the vaccinated group had higher titers across their first two years of life (**Fig 3A**), remaining elevated above the natural decay baseline of unexposed infants (**Fig S9**).

**Figure 3.**
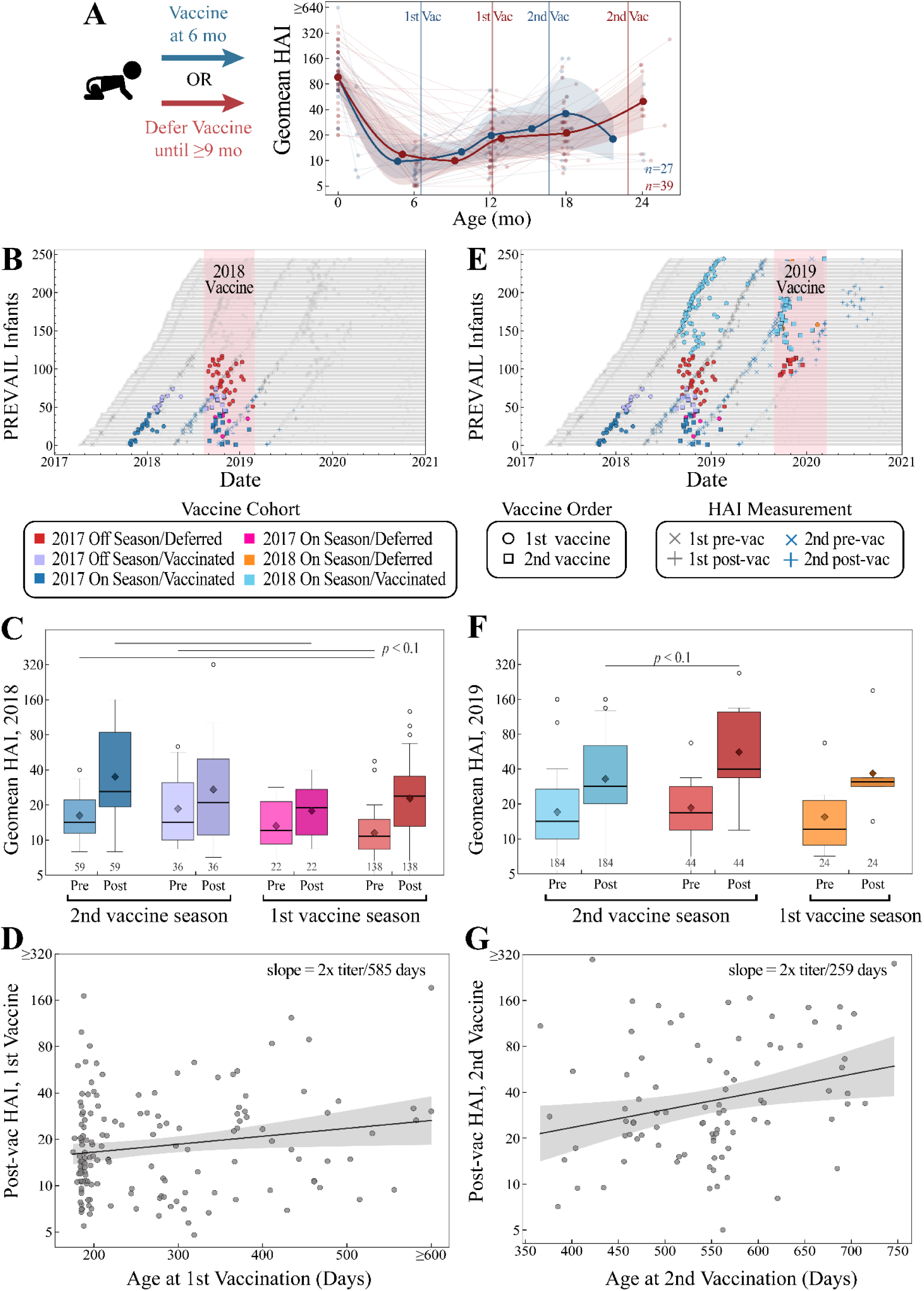
Vaccine-elicited antibody titers in infants that deferred vaccination or received it when they turned 6 months old. (A) Summary of HAI titer trajectories when infants are vaccinated at 6 months vs 9 or more months. Vertical lines represent the mean 1st and 2nd vaccination age for each group. (B-G) Infants were categorized based on if they turned 6 months old and were eligible for vaccination in July-January (*on-season*) or February-June (*off-season*), and if they received their 1st vaccine within the same season they turned 6 months old (*vaccinated*) or after that season ended (*deferred*). For each group, child geomean HAI titers across all four vaccine strains were evaluated in the (B-D) 2018 or (E-G) 2019 season. (B, E) Overview plots of vaccinations and HAI titer measurements for each group. (C, F) Box plots showing pre- and post-vaccination titers of 2018 and 2019 season, respectively. All box plots show the interquartile range, horizontal lines the medians, and dark diamonds the geometric mean titers (GMTs). Whiskers extend to 1.5 times the interquartile range. The number of measurements=(# of infants)×(# of vaccine strains) are shown for each box plot. (D) Post-1st-vaccination and (G) post-2nd-vaccination titers vs child age during 1st vaccination. *: *p* ≤ 0.05 (Welch’s t-test on the log-transformed values).

To fully assess the impact of late-season vaccination in the northern hemisphere, infants were categorized as *on-season* (born January-July, turned 6 months old in July-January) or *off-season* (born August-December, turned 6 months old in February-June), and then further split into *vaccinated* (received their 1st vaccine in the same season they turned 6 months) or *deferred* (received their 1st vaccine after the season they turned 6 months old).

We first categorized infants based on their date of birth in 2017 and compared their responses to the 2018 vaccine ‒ representing the 1st vaccination for the deferred and 2nd vaccination for the vaccinated groups ‒ using their geomean titer across all vaccine strains (**Fig 3B**). Both the on-season vaccinated (**Fig 3C, Fig S10A**, blue) and off-season vaccinated (violet) groups started with ∼1.5x higher HAI titers pre-vaccination and exhibited ∼1.6x higher titers post-vaccination than the deferred groups (pink/red). Comparing the combined vaccinated groups versus the combined deferred groups led to higher post-vaccination titers (*p*=0.129, Welch’s t-test, **Fig S11A**). Thus, receiving the late-season 1st vaccination mildly enhanced pre-and post-vaccination titers during the subsequent season.

Using these groups of infants born in 2017, and two additional groups categorized by their date of birth in 2018 (**Fig 3E**, light blue/orange), we assessed responses to the 2019 vaccine. Despite updates to both the H1N1 and H3N2 vaccine strains from 2018 to 2019 (**Table 3**), infants born in the 2018 season or their counterparts born in the 2017 season exhibited similar pre- and post-2nd-vaccination titers (**Fig 3F** light blue vs **Fig 3C** blue, **Fig 3F** orange vs **Fig 3C** red).

**Table 3.** Vaccine information for the influenza seasons spanning the PREVAIL cohort. 1st-2nd columns: North hemisphere vaccine strains over each season analyzed. 3rd column: Number of HA mutations between each vaccine strain and the prior season’s vaccine strain.

| Vaccine Season | Recommended Vaccine Strains (Northern Hemisphere) | HA Amino Acid Changes from Prior Vaccine |
| --- | --- | --- |
| 2019 | H1N1 A/Brisbane/2/2018 | 8 |
|  | H3N2 A/Kansas/14/2017 | 16 |
|  | B Victoria B/Colorado/6/2017 | 0 |
|  | B Yamagata B/Phuket/3073/2013 | 0 |
| 2018 | H1N1 A/Michigan/45/2015 | 0 |
|  | H3N2 A/Singapore/INFIMH-160019/2016 | 6 |
|  | B Victoria B/Colorado/6/2017 | 7 |
|  | B Yamagata B/Phuket/3073/2013 | 0 |
| 2017 | H1N1 A/Michigan/45/2015 | 18 |
|  | H3N2 A/Hong Kong/4801/2014 | 0 |
|  | B Victoria B/Brisbane/60/2008 | 0 |
|  | B Yamagata B/Phuket/3073/2013 | 0 |

Surprisingly, the major difference across the 2019 vaccine responses emerged between the 2017 off-season deferred infants (**Fig 3F**, red) and 2018 on-season vaccinated (**Fig 3F**, light blue). While both groups had similar pre-vaccination titers, the off-season deferred group had higher post-vaccination titers (*p*=0.094, Welch’s t-test). Both groups received exactly one prior vaccine, and since there was no difference between on- and off-season infant 2018 responses (**Fig S11B**), age was a plausible distinguishing factor. We therefore assessed whether age at vaccination was associated with the 1st and 2nd vaccine responses across all infants.

Across all infants, there was a weak positive association between infant age and post-vaccination titers following their 1st vaccine (**Fig 3D**, *Pearson r*=0.18). In contrast, there was a stronger association and an overall increase in titers with increasing age at 2nd vaccination, corresponding to an approximately 2x increase in post-2nd-vaccination titers for every 259 day increase in age at 2nd vaccination (**Fig 3G**, *Pearson r*=0.26). An even stronger positive association was observed when post-2nd-vaccination titers were analyzed as a function of age at 1st vaccination (*Pearson r*=0.40). Consistent with this, children receiving their 1st vaccine when they were 12 months old had ∼2x higher post-2nd-vaccination titers than infants that received their 1st vaccine when 6 months old (**Fig 3F**). Notably, while the strongest post-2nd-vaccination response came from late-season infants deferring their 1st vaccination, this enhancement came at the cost of these children having lower titers at ∼6-12 months of age followed by consistently lower titers at 12-24 months, until they received their 2nd vaccination (**Fig 3A**).

### 6) High-Resolution Time Dynamics of Child Vaccine Responses Revealed a Dominant Responder and Subdominant Non-Responder Phenotype

Whereas in a vaccine study all participants are measured at the same pre- and post-vaccination time points, the longitudinal nature of the PREVAIL cohort meant that child titers were measured at a nearly continuous range of times, enabling a detailed breakdown of the dynamics following the 1st and 2nd vaccination. HAI trajectories were calculated using geomean titers across rolling time windows, spanning from the pre-vaccination baseline to 300-400 days post-vaccination (**Methods**). Titers measured after an influenza infection were excluded from the homologous subtype/lineage, and the geometric mean was calculated using the remaining vaccine strains.

Across all children and vaccine strains, the 1st vaccine response elicited a fold-change of 1.5x around day 100 (**Fig 4A**, dashed black), comparable to the earlier time-independent analysis (**Fig 2B**, left). Yet individual trajectories revealed distinct immune phenotypes. The most prominent *responder* phenotype, defined by having increasing titers pre- to post-vaccination, showed a steady increase in titers peaking between days 100-200 (2.5x fold-change), followed by a slight waning by day 365 post-1st-vaccination (1.9x fold-change relative to pre-vaccination) (*n*=94 using all vaccine strains; **Fig 4A**, gold). Few children maintained a strong and durable response. Only 2/46=4.3% children with titers measured after day 250 post-1st-vaccination had ≥4x fold-change compared to their day 0 values, while 35/46=76.1% were within 2x of day 0 titers.

**Figure 4.**
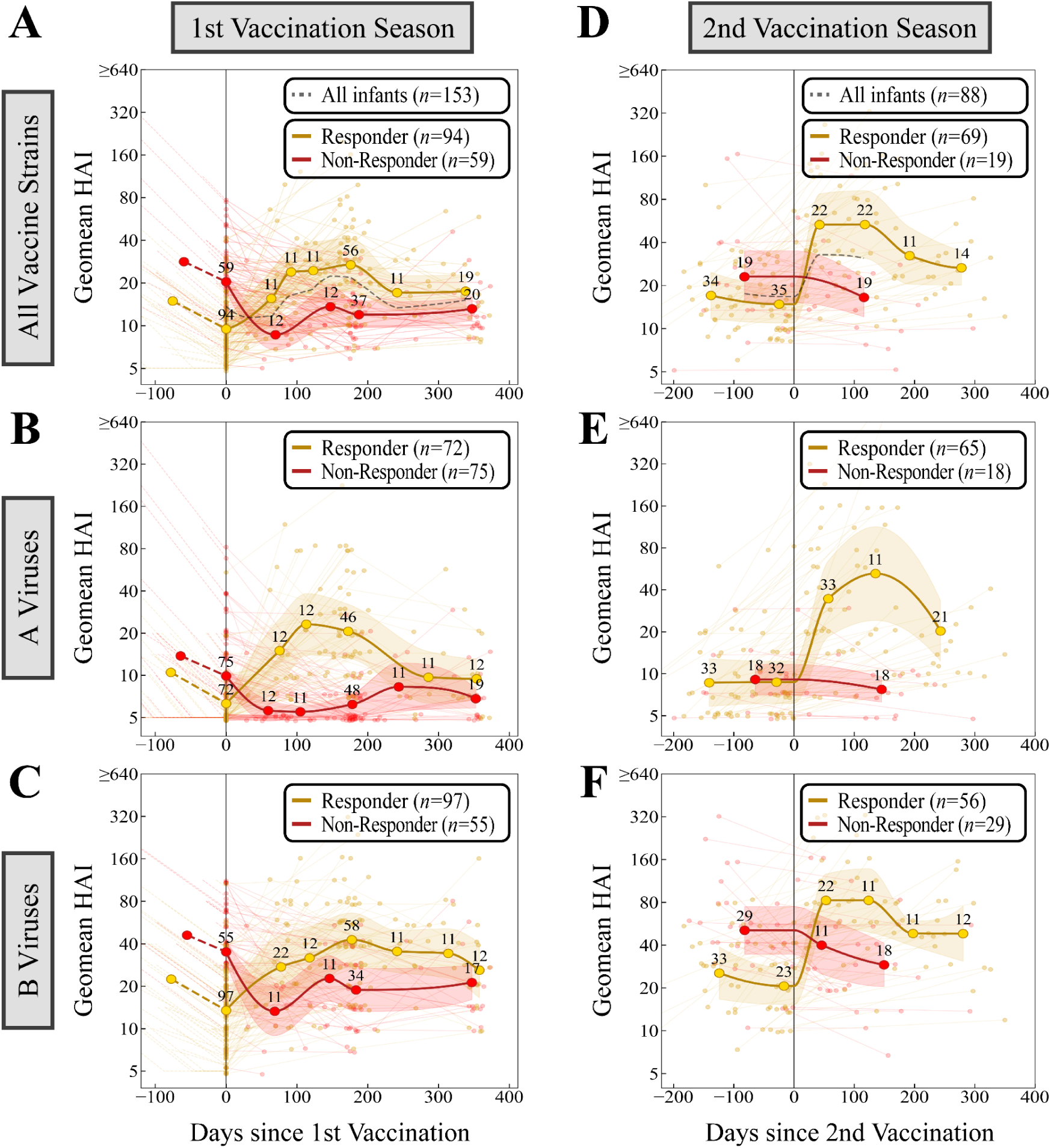
High-resolution dynamics of the 1st and 2nd influenza vaccine response in children. HAI trajectories for the responder and non-responder receiving their (A-C) 1st or (D-F) 2nd vaccination. Trajectories show the geomean titers against (A,D) all vaccine strains, (B,E) both influenza A vaccine strains, or (C,F) both influenza B vaccine strains. Time points were binned to include ≥10 children and connected by splines, with the precise number shown above each point (**Methods**). Responders (titers increase post-vaccination) and non-responders (titers decrease or stay the same) were calculated independently in every panel. Since titers to a subtype or lineage are ignored after homologous infection, the total number of A/B virus responses may be smaller when children are infected with both influenza A/B vaccine strains (**Methods**).

In the remaining *non-responder* children, titers either stayed the same or decreased post-vaccination (*n*=59 using all vaccine strains; **Fig 4A**, red). Subsetting non-responder HAI titers against influenza A or influenza B viruses revealed that the vaccine did not take for influenza A strains, with titers showing the same decay seen from maternal antibody waning (titer=10 at day 0, titer≈5 from day 50 onwards; **Fig 4B**, red). In contrast, their influenza B responses started from titer=40 at day 0, decreased by 2x, and then remained stable out to day 365, suggesting that durable antibodies were generated against B viruses or that influenza B waning plateaus at titer≈20 (**Fig 4C**, red).

Notably, non-responders comprised 75/147=51% of the cohort for influenza A viruses, far larger than the 55/152=36% seen for influenza B (**Fig 4B,C**). Further subsetting by subtype, 65% of children were non-responders for H1N1 and 52% were non-responders for H3N2 (**Fig S12A**). Non-responder groups for both influenza A and B started with larger titers at day 0, so that having higher titers on the day of 1st vaccination significantly increased the odds of a worse vaccine response (*p*≤0.05, Welch’s t-test), likely reflecting maternal antibody inference (**Fig S13**).

After the 2nd vaccination, the responder phenotypes showed a more rapid response, with titers increasing immediately after day 0 and reaching their peak response around day 40 (**Fig 4D**). These peak titers were maintained until day 120 and then declined. Strong and durable immunity was slightly more common. Of the children with titers measured after day 250 post-2nd-vaccination, 2/12=16.7% of children had ≥4x fold-change compared to their day 0 values, while 5/12=41.7% were within 2x fold-change.

In contrast, non-responders remained quite stable for their entire 2nd vaccine response, staying within 2x of their day 0 titers and either mildly decreasing around day 150 by 1.2x against influenza A viruses or decreasing by 1.7x against influenza B viruses. Notably, children who were influenza B non-responders during the 1st vaccination generally did not remain influenza B non-responders during the 2nd vaccination, which is why the former group had titers≈20 at day 365 post-1st-vaccination but the latter group had titers≈40 pre-2nd-vaccination. Moreover, while influenza B non-responders had higher titers than responders pre-2nd-vaccination, the influenza A responders and non-responders had similar pre-2nd-vaccination titers, suggesting that other factors dictate the phenotype of the 2nd vaccine response to influenza A.

Collectively, the 1st vaccine response elicited a small but permanent increase in antibody titers, whereas the 2nd vaccine response elicited a faster and stronger response where titers decayed back towards baseline. Influenza B titers were consistently elevated compared to influenza A titers at all time points and for both responder and non-responder phenotypes.

### 7) Pre-vaccination serology was the most accurate determinant of post-vaccination titers

A notable trend of adult influenza antibody titers is their heterogeneity, which is often ascribed to an individual’s complex and often-unknown prior infection and vaccination history. Yet the PREVAIL infants also displayed heterogeneous 1st and 2nd vaccine responses ranging from fold-change≤1x (vaccine did not take) to fold-change=256x. Given each infant’s known vaccination history and the lack of prior influenza infections, we assessed what features affected their antibody titers post-vaccination.

Despite capturing a rich set of maternal and infant features (*e.g.*, sex, maternal age at delivery, maternal smoking, infant gestational age at birth, delivery type, duration of breastfeeding), only infant sex showed a noticeable effect, while none of the other features substantially impacted post-vaccination titers (**Fig S14**). When comparing post-1st-vaccination titers and fold-change, female children demonstrated slightly stronger responses across all strains compared to males. This sex-based divergence was particularly evident when the dataset was subset to infants with low pre-1st-vaccination titers (**Fig S14A**). While these features offered little predictive value, an infant’s longitudinal HAI trajectory emerged as the best predictor of their post-vaccination response.

For the 1st vaccination, children whose maternal antibodies had not fully waned had worse responses. More precisely, children with an imputed HAI titer≥10 for influenza A or ≥20 for influenza B on the date of their 1st vaccination were more likely to elicit no response and have fold-change≤1x (94%=31/33 children for H1N1, 84%=41/49 for H3N2, 72%=41/57 for B Victoria, 59%=51/87 for B Yamagata). In contrast, children with lower titers pre-1st-vaccination had far smaller odds of having fold-change≤1x (57%=60/106 children for H1N1, 35%=31/89 for H3N2, 25%=23/94 for B Victoria, 19%=12/63 for B Yamagata). Similar statistics hold without imputation by only considering infants with HAI titers measured within 20 days of 1st vaccination. As noted above, H1N1 was the only vaccine component that did not take in the majority of PREVAIL infants, yet the likelihood of not responding substantially increased when pre-vaccination titers were ≥10.

The 2nd vaccine response for influenza A could be partially predicted based on a child’s 1st vaccine response. A moderate 1st vaccine response (fold-change≥2x) led to a moderate 2nd vaccine response (fold-change≥2x) in 81%=13/16 infants for H1N1 and 81%=26/32 infants for H3N2, while holding more weakly across 49%=21/43 of infants for B Victoria and 46%=15/33 for B Yamagata. Moreover, cases where maternal antibodies were appreciable and the 1st vaccine did not take (fold-change<1x) led to no 2nd vaccine response (fold-change≤1x) in 68%=15/22 of infants for H1N1, although this trend only held for a minority of infants for H3N2, B Victoria, or B Yamagata. Ultimately, these results demonstrate that the heterogeneity in these 1st and 2nd vaccine responses are partly attributable to each child’s prior serological trajectory.

### 8) Influenza A infections elicited a subtype-specific antibody response that was stronger, more rapid, and more durable than vaccination

Influenza infections are expected to elicit robust antibody responses that are comparable to, or exceed, those induced by vaccination. Within the PREVAIL cohort, nasal swabs specified the subtype (*n*_H1N1_=19 or *n*_H3N2_=16) or lineage (*n*_B Vic_=6 or *n*_B Yam_=2) of the infecting strain. Pre- and post-infection titers demonstrated that influenza A infections were subtype-specific, with H1N1 not impacting H3N2 titers and vice versa (**Fig 5A, Fig S15**).

**Figure 5.**
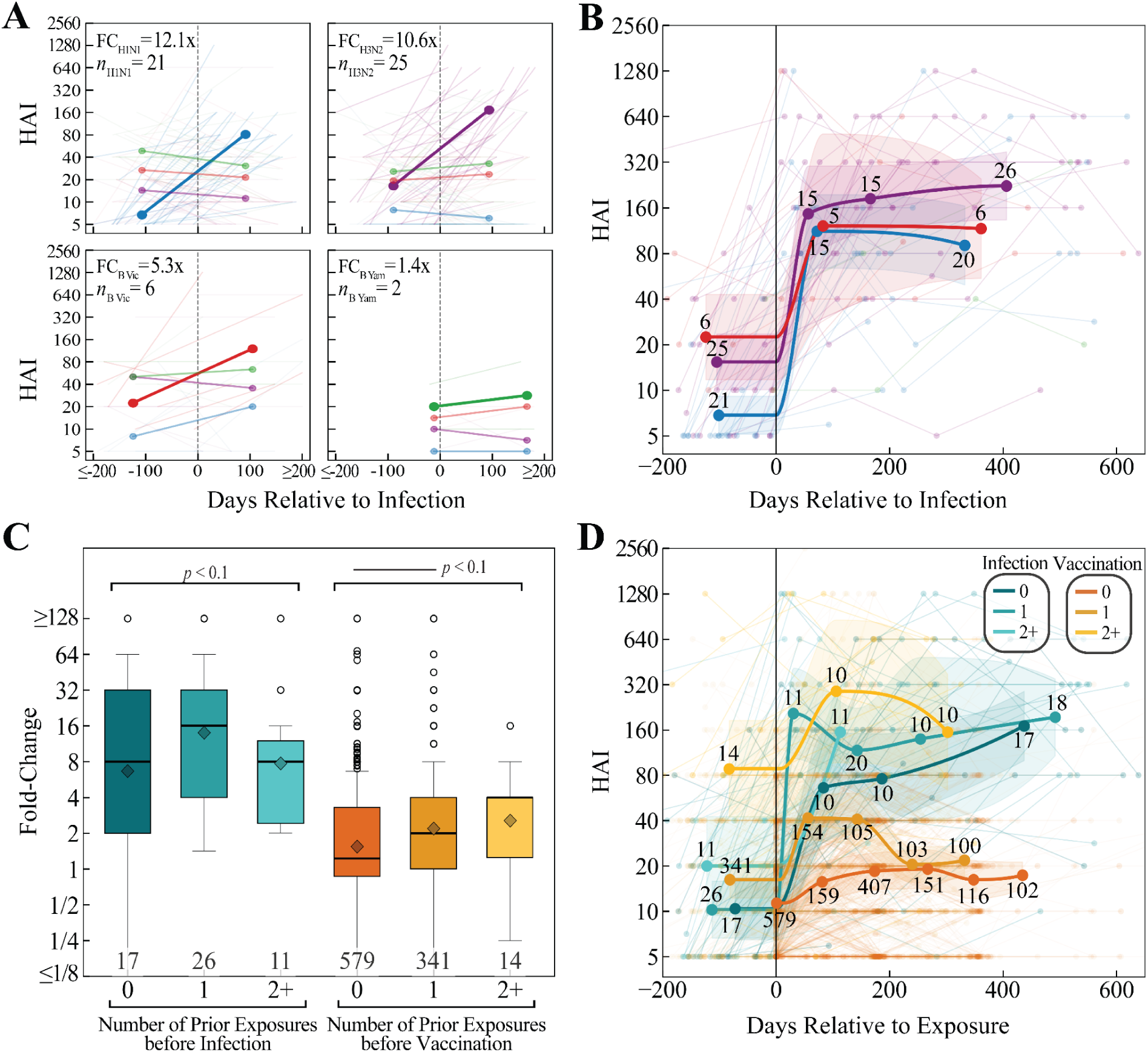
Impact of Infections on the antibody response. (A) Pre- and post-infection HAI titers for each influenza subtype or lineage. Individual trajectories are shown in thin lines, with geometric mean titers represented by bold lines. The infecting subtype/lineage is highlighted with a thicker line. (B) Longitudinal dynamics of HAI titers following infection for four vaccine components. (C) Comparisons of HAI fold-change across groups stratified by exposure history [exposure=infection or vaccination (one or two doses) in one season]. (D) Longitudinal dynamics of HAI titers for each exposure group. In Panels B and D, time points were binned to include ≥5 children and connected by splines, with the precise number shown above each point (**Methods**). Bracket indicates significance against all other groups. *: *p* ≤ 0.05 (Welch’s t-test on the log-transformed values).

Infections were also identified through medical records where rapid influenza diagnostic tests were administered that only specified if the infecting strain was influenza type A (*n*_A_=14) or type B (*n*_B_=3). Of the 17 cases where a nasal swab and medical records identified an infection within 3 weeks of each other, the subtype and type always matched, confirming the consistency between both detection methods. Using subtype-specific serological responses as a defining feature, 11/14 of the influenza A infections were confidently categorized as H1N1 (*n*_H1N1_=2) or H3N2 (*n*_H3N2_=9), while the remaining 3 infections showed similar titer increases to both H1N1 and H3N2; these final cases could not be categorized and were removed from the subsequent analyses (**Fig S15B**).

H1N1 and H3N2 infections elicited a substantial boost in HAI titers, resulting in a fold-change of 12.1x and 10.6x, respectively, using the nearest pre- and post-infection time point (**Fig 5A, Fig S15E**). This HAI boost was highly specific to the infecting subtype, and no comparable increases were observed in the other vaccine components within the same individual. Furthermore, fold-change ≥4x occurred in 15/21 H1N1 and 20/25 H3N2 infections, underscoring the potency of the antibody response to influenza A infection.

In contrast, there were far fewer B Victoria and B Yamagata infections. Among these cases, B Victoria infection elicited 5.3x fold-change while B Yamagata elicited a small 1.4x fold-change pre- to post-infection (**Fig 5A**). In addition, the four influenza B infections identified through medical records showed increasing titers to both B Victoria and B Yamagata, and hence the infecting strain’s lineage could not be inferred (**Fig S15D**). Among the eight nasal swab cases with known lineage, the fraction of infections with fold-change ≥4x was 3/6 for B Victoria and 0/2 for B Yamagata. While there was a post-infection antibody boost from influenza B infections, it was far less pronounced than the response from influenza A infection.

The longitudinal dynamics from influenza A infections showed that the antibody response was rapid and highly durable. H1N1 and H3N2 titers peaked around day 50-70, and these titers remained elevated for at least 330 days post-infection (**Fig 5B**, blue and purple), a stark contrast to the low HAI titers of unexposed children (**Fig S9**). Based on six B Victoria infections, that lineage’s titer dynamics showed an equally rapid response comparable to H3N2 dynamics, although the B Victoria pre-infection titers were larger due to the higher natural baseline of B Victoria (**Fig 5B**, red, **Fig S9**). The limited number of B Yamagata cases (*n*_Yam_=2) was insufficient to infer those dynamics.

To assess the impact of exposure history (*i.e.*, the number of prior infections and vaccinations, regardless of their order), HAI fold-change was quantified for infants with 0, 1, or 2+ prior exposures followed by either an infection or vaccination. Infection responses were far stronger than vaccination responses regardless of prior exposures, but also showed far more variability, and there was no clear trend between the number of prior exposures and infection-elicited fold-change (**Fig 5C**, teal groups). For vaccinations, there was a small but steady increase in fold-change with the number of prior exposures. Notably, the *n*=14 children with 2+ prior exposures followed by vaccination showed the highest pre-vaccination titers, since this group must have received at least one prior infection, yet they still achieved slightly larger fold-change than children with fewer prior exposures (**Fig 5D**). Taken together, these results suggest that prior infection in children led to durably higher baseline titers, yet both vaccinations and infections have modest effects on the fold-change of subsequent exposures.

Lastly, we assessed whether symptomatic infections led to greater fold-change compared with asymptomatic infections. Two methods were used to categorize infections as symptomatic: 1) the infant had a medical visit with reported influenza-related symptoms – fever, cough, vomiting, diarrhea, dehydration, or altered mental status – within ±10 days of the infection-positive nasal swab. 2) The infection-positive nasal swab occurred during the duration of an acute respiratory infection episode reported by the mother via weekly text message. No significant differences were observed in fold-change between symptomatic and asymptomatic infections using either method, suggesting that symptom severity did not correlate with the magnitude of the antibody response (**Fig S16**).

## Discussion

Since infant cord blood titers closely matched maternal serum titers during delivery and then waned over time, increasing a pregnant mother’s influenza titers provides a stronger and more durable antibody defense to the infant during their first months of life. While this protects infants when they are most vulnerable, it can also blunt their own initial vaccine response. Mothers vaccinated during the 2nd or 3rd trimesters (but at least 30 days before delivery) led to a small increase in titers compared to earlier or later vaccination, corroborating past observations for influenza or TDaP^23–25^ and consistent with vaccine-elicited adult antibody kinetics where the peak antibody response is reached 10-75 days post-vaccination before waning.^5^ Surprisingly, unvaccinated mothers had significantly lower titers than mothers vaccinated 12-18 months pre-delivery, even though antibody titers in this latter group should have returned to baseline in most individuals by one year post-vaccination.^5^ Future work should determine whether the onset of pregnancy alters these antibody kinetics.

Mothers <25 y.o. during delivery had 2x larger titers than mothers >35 y.o., matching the observed trends across thousands of male and female vaccinees across 10+ influenza seasons, suggesting that this age-based trend is not altered by pregnancy.^17^ However, rotavirus IgG titers were measured for these same mothers, but anti-rotavirus IgG titers did not decrease with increasing maternal age.^26^ Thus, the 2x decline in titers between age 25-35 could be an influenza-specific phenomenon.

H1N1, H3N2, B Victoria, and B Yamagata titers decayed exponentially, irrespective of the cord blood titer levels. For example, an infant with HAI_H1N1_=640 during delivery would maintain HAI_H1N1_≥40 for ∼5 months longer than an infant born with HAI_H1N1_=80. Hence, any boost in cord blood titers from maternal vaccination or age are maintained throughout the first months of life, as seen in prior studies.^14^

The waning of maternal antibodies provides a unique setting to measure antibody half-life in the absence of continual antibody generation. Curiously, influenza B antibodies exhibited significantly longer half-lives and thus more durability than influenza A antibodies. This signal was highly robust, as seen for all ∼221 infants with ≥3 HAI titer measurements before their first exposure. While we did not investigate the biochemical mechanism underlying these different half-lives, prior work has shown that enhanced antibody affinity to the neonatal Fc receptor or changes in glycosylation can lead to increased half-life.^27–30^ While many antibodies show a half-life of ≲40 days in humans,^31,32^ therapeutic antibodies with Fc modifications reached half-lives of 70-100 days,^33–35^ in line with the influenza B titer half-lives, and thus we speculate that the Fc regions of B Victoria and B Yamagata antibodies may facilitate their increased half-life. Due to this slower decay, infants had higher influenza B titers pre-1st-vaccination, and influenza B titers continued to exceed those of influenza A post-1st-vaccination as well as pre-and post-2nd-vaccination.

This unique influenza B virus behavior is especially noteworthy in light of B Yamagata’s extinction in 2021.^36,37^ Recent work has shown that influenza B has less within-host diversity^38^ and is more susceptible to higher temperatures.^39^ A compilation of large-scale serological studies revealed that B Yamagata titers persisted for at least two years after B Yamagata became extinct in 2021,^17^ potentially due to the slow genetic and antigenic evolution of that lineage. The differences in half-life revealed in these infants may have also contributed to this lineage’s extinction, particularly within the context of epidemiologic changes in influenza transmission and incidence during the COVID-19 pandemic.^40^

Because of the exponential decay in influenza titers, imputation was critical when quantifying the 1st vaccine response in the PREVAIL cohort, since some children were sampled weeks or months prior to vaccination. Even with imputation, the 1st vaccination elicited a small GMT fold-change of 1.3-1.5x across the entire cohort for each vaccine strain.

Prior reports on responses of infants and children have greatly varied. One 2008 study of children 0.5-3 y.o. receiving their 1st vaccine (two full doses of Fluarix) reported substantial fold-change with pre→post-vac GMTs of 8.7→82.3 (H1N1), 10.8→113.1 (H3N2), and 8.5→182.6 (Yam) (fold-change range: 9.5-21.5x).^41^ Another set of children in this same age group receiving a comparator vaccine (two *half* doses of Fluzone) showed stronger responses of 8.6→191.3 (H1N1), 11.2→247.7 (H3N2), and 8.2→232.6 (Yam) (fold-change range: 22.1-28.4x).^41^ Other work has ranged from reporting equally large antibody responses^42^ to surprisingly small responses where only 2/7 infants showed any measurable neutralization titers above baseline against a panel of H3N2 viruses.^43^ Prior work has also suggested that influenza B vaccine strains elicited the weakest antibody response,^12,13^ showing the opposite behavior to what was seen in PREVAIL. Given that the primary endpoints in most influenza vaccine trials is to elicit large HAI titers and large fold-change, these collective results highlight a pressing need to understand this vast discrepancy in the antibody response elicited by some antigens but not others.

Importantly, these discrepancies may be driven by the fundamental differences between controlled clinical trials and real-world observational cohorts. While clinical trials strictly enforce two-dose vaccine schedules at precise intervals to maximize immunogenicity, the PREVAIL cohort reflects real-world variability in vaccine timing and adherence. Furthermore, the immunological landscape has shifted significantly since these earlier studies were conducted, with most historical cohorts involving infants born to mothers with little-to-no pregnancy-related influenza vaccination. In contrast, the high rate of maternal vaccination in the PREVAIL cohort (83%) likely resulted in higher baseline maternal antibodies, which could alter infant vaccine responses.

PREVAIL infants receiving their 1st vaccine had an unusually high likelihood of not responding to the H1N1 vaccine strain (65% had fold-change_H1N1_≤1x). Roughly 60-70% of adults receiving influenza vaccination consistently show fold-change≤1x for H1N1 and H3N2, presumably due to prior exposure and high pre-vaccination levels.^5^ Yet PREVAIL children had no prior exposures, had low pre-vaccination titers, and >50% of them exhibited fold-change≥2x against the H3N2, B Victoria, and B Yamagata vaccine strains. In short, the H1N1 component did not take in a majority of these infants, in stark contrast to the other vaccine strains, so that even vaccinated children remained susceptible to H1N1 infections.

Higher pre-1st-vaccination titers against any vaccine strain increased the likelihood of an infant not responding to that strain. This suggests that pre-existing maternal antibodies can interfere with the vaccine response, as noted in earlier studies.^13,44^ For example, in a 2004 vaccine study administering two half-doses to infants 6-12 weeks or 24-36 weeks old, the older infants had 1.3-4.2x smaller titers pre-vaccination but achieved 1.7-2.0x larger titers post-vaccination.^14^ This suggests that the 1st vaccination should be administered after an infant’s titers fall to HAI=5 across all vaccine strains. Whereas prior studies, many conducted more than a decade ago, found that most 6 months old infants had HAI titers <10 for each vaccine strain,^12–14^ only 6% of PREVAIL infants achieved this limit for all strains at 6 months old. This incomplete decay was primarily due to the longer half-lives of B Victoria and B Yamagata antibodies, which may be a recent phenomenon. Yet these results suggest that maternal antibody interference may be currently occurring in a majority of infants for at least one influenza vaccine strain.

To that end, the time when each infant reaches undetectable antibody titers for all vaccine strains can be readily determined from maternal serum titers at delivery and the known half-lives of each vaccine strain. Based on antibody decay kinetics, most PREVAIL children would have achieved this limit when they were 13 months old (range 5-21 months old; 9 months old for influenza A [range 0-16 months] and 12 months for influenza B [range 2-21 months]). Such methods present an opportunity to optimize vaccine administration on a per-person basis, and since a stronger 1st vaccine response led to a greater likelihood of a stronger 2nd vaccine response, such approaches could provide the foundation for better long-term immunity.

Among PREVAIL children, antibody fold-change after the 2nd vaccination was slightly larger for influenza B and substantially larger for influenza A, with the H1N1 GMT fold change of 2.7x approaching the ∼3x fold-change generally seen in adults for all vaccine strains.^17^ Whereas the 1st vaccine elicited a small antibody response that only mildly diminished by 365 days post-vaccination, the peak response after 2nd vaccination lasted for ∼150 days, nearly twice as long as the peak response in adults.^5^ Thus, children exhibit fundamentally different 1st and 2nd vaccine kinetics, yet they rapidly converge towards adult-like responses in their first years of life.

Unlike adults, infants can only receive an influenza vaccination once they are ≥6 months old, raising the question of whether infants that turn 6 months in February-May in the northern hemisphere should receive a vaccination or wait until the next season. Current influenza vaccination policies are complex, but they do not offer a concrete suggestion for this situation.^22^ PREVAIL findings provide a clear immunological rationale for late-season vaccination: the 1st vaccination acts as a stable, long-term primer that, despite low initial titers, augments the 2nd vaccination response. In contrast, infants turning 6 months in February-May that abstained from late-season vaccination had very low antibody titers during the start of the next influenza season and continued to have low titers for a year after their 1st vaccination. Interestingly, if such infants were not infected with influenza, they showed the strongest fold-change after their 2nd vaccination. Future work will follow these same infants into additional seasons to determine the long term impact of deferring or accepting late-season vaccination.

Compared to vaccination, pediatric infection responses remain understudied. Among the 245 PREVAIL children, 60 influenza infections were detected during their first two years of life. We found that antibody responses to influenza A infections were substantially stronger than vaccine-elicited responses and highly subtype-specific. Other work has shown that influenza A infections can elicit exceptionally broad responses within the same subtype, where 7/15 infants infected with pre-pandemic H1N1 in 2007 or 2008 exhibited measurable titers against the antigenically distinct pandemic H1N1 A/California/7/2009.^45^ Responses to influenza B infection in PREVAIL were weaker than those from influenza A infections, but they showed some cross-reactivity between B Victoria and B Yamagata. Prior work using a Hong Kong cohort comprised primarily of children 6-17 y.o. found that in half of the infection cases, subjects had HAI=5 both pre- and post-infection against the lineage of infection.^46^ Among subjects with an HAI fold-rise, B Victoria or B Yamagata infection led to a fold-rise of 4.4-5.7x in the infecting lineage and ∼1/3 of this fold-rise in the other lineage.^46^ These data suggest that influenza A infection yields a stronger, subtype-specific antibody response while influenza B elicits more modest but cross-reactive immunity.

One limitation of this study was that it analyzed a single infant cohort. While this ensures a consistent study design and HAI assay administration, it is unclear how these results would generalize to other populations or other influenza seasons. Although most infants received two doses 1-2 months apart during their 1st vaccination season, the kinetics following each dose could not be examined since sera were collected 6 months apart. In addition, the exponential decay of maternal antibodies (linear on a log scale) assumed a constant decay rate until an undetectable HAI titer=5 was reached. If influenza B viruses have a higher minimum titer, their fold-change may have been over-estimated. All of these features are worth exploring in additional datasets.

Vaccinating infants for influenza is crucial, as 85% of pediatric deaths reported to CDC during the 2025-26 season occurred in children who were not fully vaccinated against influenza.^47^ The PREVAIL cohort sheds light on how influenza vaccines can be further improved by demonstrating that the “clean slate” of infant immunity is less a blank canvas and more a complex stage set by maternal antibody transfer. Understanding the differences in influenza A versus influenza B responses and the vast heterogeneity reported across different seasons will be essential for designing next-generation pediatric vaccines that can bypass maternal antibody interference, provide durable immunity, and pave the road for strong subsequent vaccine responses.

## Methods

### Study design

Pregnant women residing in the Cincinnati metropolitan area, 18 years and older, were invited to participate in the Pediatric Respiratory and Enteric Virus Acquisition and Immunogenesis Longitudinal (PREVAIL) cohort. Mothers with a live-born, healthy, singleton pregnancy of at least 34 weeks of gestation, that provided written consent and completed their first postnatal study visit at week 2 were officially enrolled. Further details on the study design, inclusion/exclusion criteria, and data collection have been described previously.^48^ This study was reviewed and approved by the institutional review boards at the Centers for Disease Control and Prevention (CDC), Cincinnati Children’s Hospital Medical Center (CCHMC), and the enrolling birth hospitals: The Christ Hospital and University of Cincinnati Medical Center.

Substantial effort was put into tracking infections and vaccinations. Infections were assessed through weekly nasal swabs with influenza RT-PCR that were screened regardless of symptoms, as well as rapid influenza diagnostic tests performed during medical visits. Despite this intensive survey, some infections may not have been detected, especially if a nasal swab was missed or if a mild infection was cleared during the interval between nasal swab submissions. Influenza vaccine administration data for both mother and infant were captured and validated through state vaccine registries and medical records.

### Influenza viruses

All viruses were propagated in eggs and obtained through the International Reagent Resource (IRR), Influenza Division, World Health Organization Collaborating Center for Surveillance, Epidemiology and Control of Influenza, CDC, Atlanta, Georgia.

### HAI protocol

HAI quantified the ability of serum antibodies to inhibit viral hemagglutination of red blood cells. Reference antigens were titered to get their HA unit each day the assay was performed, with 4 HA units/25 µL of viral antigen added to 2-fold dilutions of sera. Each serum sample was first treated with receptor destroying enzyme (RDE) to remove nonspecific inhibitors. A 1:4 dilution of sera to RDE solution was incubated in a 37°C water bath for 18–20 hours, then transferred to a 56°C water bath for 30-60 minutes to inactivate the RDE.

Turkey red blood cells (RBCs) from a qualified source were used. After washing, 0.5 mL of the packed, buffy-coat free RBCs were added to 100 mL PBS to create a 0.5% suspension of cells. To prevent non-specific binding, the RDE-treated sample sera were absorbed with packed RBCs to reach a final dilution of 1:20. This mixture was incubated at 4°C for one hour, with mixing every fifteen minutes, and then centrifuged to remove the adsorbed serum without disturbing the cells.

Each serum sample was tested in duplicate, starting at 1:10 with a two-fold dilution series in PBS leading to a range of 10-2560 dilution in V-bottom microtiter plates. Serum-only and RBC-only control wells were included on each plate. Starting dilutions were adjusted for higher titer samples. Equal volumes of antigen (4 HA units/25 µL) to sample were added in each well, mixed thoroughly and incubated at room temperature for 30 minutes. Equal volumes of 0.5% washed RBCs were added to each well of the plate, mixed and incubated at room temperature for 30 minutes. Plates were read visually by tilting and documenting the last well completely inhibiting hemagglutination. The HAI titer is the reciprocal of the last dilution of serum that completely inhibits hemagglutination. Titers less than 10 were reported as an HAI titer=5.

The duplicate titers for each sample must have been within 2-fold of each other or the sample was retested. If titers differed by ≤2-fold the lower of the two titers was used. The assay was considered valid if both the high and low control sera were within 2-fold of their expected titers and the RBC control wells were completely negative.

As in prior work, titers were analyzed on a log scale but plots were reported in the original titer units. Since HAI takes on discrete values, points were jittered for clarity when necessary. Unless otherwise stated, each analysis included all eligible participants; for example, the maternal-cord blood analysis considered all maternal-dyad pairs with available measurements, regardless of the date of delivery or the timing of maternal vaccination.

### Defining 1st and 2nd vaccinations as well as infections

The 1st influenza vaccine dose a child received, regardless of their age or if a second dose was received that season, was defined as their date of 1st vaccination. Additional influenza vaccinations given during that same season (July 1 through the following June 30), regardless of their timing, were counted as multiple doses for this same 1st vaccine season. The 2nd influenza vaccine was similarly defined as the next vaccine administered in any following influenza season, with additional vaccinations during that season counting as multiple doses.

Serum was drawn based on child age (at 0, 1.5, 6, 12, 18, and 24 months) and not aligned to vaccinations, and in some current and/or prior vaccine strains were measured by HAI. Pre- and post-vaccination titers were defined as follows:

● 1st vaccine season:

○ Pre-vaccination: The latest HAI measurement either before 1st vaccination or up to 14 days post-vaccination, since the dynamics analysis showed that titers do not rise within this window.
○ Post-vaccination: The earliest HAI measurement from 28 days post-vaccination up until 7 days post-2nd-vaccination, since titers do not rise within this short window.
● 2nd vaccine season:

○ Pre-vaccination: Defined as any measurement taken from post-vaccination period of the 1st vaccination up to 7 days after the 2nd vaccination. If multiple data points existed, the most recent HAI titer was used, as high-resolution dynamics of the 1st vaccine response show little change from day 250 onward (**Fig 4A-C**).
○ Post-vaccination: Defined as the closest measurement taken more than 14 days after the 2nd vaccination, up until 7 days after the next vaccination.

When multiple strains were measured by HAI, pre- and post-vaccination titers were always selected to have the matching strain from the current season when possible. After any influenza infection, titers from the homologous infection strain (H1N1, H3N2, B Victoria, or B Yamagata) or infection type (influenza A or influenza B, if the specific strain could not be inferred) were excluded from that point onwards while retaining the titers for all other strains. For example, if an infant was infected with H3N2 before their 2nd vaccination but after their post-1st-vaccination serum collection, all of their data was added in the 1st vaccination analysis and only their H1N1, B Victoria, and B Yamagata titers were used from their 2nd vaccine response. If a child was infected before their 1st vaccination with influenza A (with unknown subtype), then only their B Victoria and B Yamagata titers would be used when analyzing the 1st and 2nd vaccine responses.

For the infection analysis, if multiple infections occurred within 28 days, the infection event with the earliest date was selected. If a specific subtype or lineage was provided, it was used, otherwise, the event was marked as influenza A or B and further classified based on its trend between pre- and post-infection measurement (**Fig S15**). If HAI titers were measured before the infection date, it was considered as pre-infection, and if it was measured on or after the infection date, it was defined as post-infection. Unlike in the vaccination analyses, pre- and post-infection time points were used irrespective of other infections or vaccinations happening between the infection-of-interest and serum collection (**Fig 5C,D**).

### Imputing HAI titers pre-1st-vaccination

The rate of maternal antibody waning was inferred for each influenza strain separately. Only infants with at least two measured titers before their 1st vaccination were included, and after any influenza infection the subsequent titers from the homologous infection strain were excluded. Lastly, if titers hit the lower level of detection (HAI=5), then all subsequent measurements were ignored for that infant and strain. With the resulting data, log_2_(HAI titers) versus infant age fit by linear regression for each vaccine strain to obtain the negative decay slope *m*_strain_ and half-life *T*_1/2_= -1/*m*.

Combining the decay rate for each vaccine strain (**Fig 2B**), inferred using all pre-1st-vaccination HAI measurements across the cohort, with measured pre-1st-vaccination titer for each infant enabled an estimation of that infant’s titers on the exact date of 1st vaccination. More precisely, the negative population-average slope (*m*_strain_) of HAI titer decay over time was fit using log-scaled titers for each vaccine strain. Given the time Δ*t* between an infant’s pre-1st-vaccination titer (HAI_pre_) and their date of 1st vaccination, titers were imputed using the first three HAI measurements (0, 1.5, and 6 months) as HAI_pre_·2*^m^*^·Δ*t*^. Imputation was only done when HAI_pre_ was measured 1 or more days before the date of vaccination and the infant was younger than 1 year of age (after which maternal titers should have decayed); any other HAI_pre_ titer was used without imputation. Titers imputed to be ≤5 were clipped to an HAI=5.

### Cohort stratification by vaccine timing

To analyze the effects of late-season vaccination, infants were stratified based on when they turned 6 months old and became eligible for their first influenza vaccine. Infants born in January to July that turned 6 months old between July and January were classified as *on season*, as this time period coincides with when seasonal influenza vaccines are available and generally administered in the northern hemisphere. All other infants were categorized as *off season*.

Infants were also categorized as *vaccinated* if they received their 1st vaccination during the same season in which they turned 6 months old, and while infants receiving their 1st vaccination in a later season were called *deferred*.

### Computational modeling of HAI trajectories

Longitudinal HAI dynamics employed a combination of dynamic binning and Piecewise Cubic Hermite Interpolating Polynomial (PCHIP) smoothing to accurately represent HAI trends while preserving the monotonicity of the data and preventing artificial oscillations between discrete time intervals.

Data from all children were aligned by the day to 1st vaccination, 2nd vaccination, or infection. Pre- and post-exposure measurements were analyzed separately to prevent blending of the distinct phases. Data were binned as finely as possible while maintaining ≥10 measurements per bin (**Fig 4**), or when data was sparser, ≥5 measurements per bin (**Fig 5**). Adjacent bins separated by fewer than 25 days were merged. A continuous trajectory passed through each bin’s log_2_(GMT) using PCHIP interpolation, with 95% confidence intervals (CI) calculated using CI_lower_ = log_2_(GMT)-(1.96·SEM) and CI_upper_ = log_2_(GMT)+(1.96·SEM), where SEM represents the standard error of the mean of log_2_(HAI). In **Fig 4A,D**, responses for all infants (black dashed curves) were calculated using a weighted average between the responder and non-responder groups (weighed by the number of infants in each group) followed by PCHIP smoothing.

### Statistical analysis

When assessing differences between different groups, Welch’s t-test on log-transformed values was employed. For longitudinal comparisons of pre- and post-exposure measurements within the same group, the paired t-test was used. RMSE between cord blood and maternal HAI was computed on the difference in their log_2_(HAI titers) (**Fig 1B**). Linear regression slope (log_2_[fold-change] vs log_2_[pre-vac HAI]) was similarly computed for each subtype (**Fig 2C**). To assess age-dependent trends in vaccine responses, simple linear regression was performed between infant age at vaccination and log_2_[post-vac GMT], and the slope and Pearson correlation coefficient *r* were reported (**Fig 3D,G**).

## Data and Code Availability

*For reviews*: All data is provided as a supplementary information file.

*When the manuscript is accepted*: All data and code will be made available at https://github.com/TalEinav/InfluenzaInfantImmunity.

## Data Availability

All data and code will be made available on Github upon publication.

## Acknowledgements

We thank Aaron Lane for useful discussions. We gratefully acknowledge the participation and dedication of the PREVAIL cohort families. This research was supported by LJI & Kyowa Kirin, Inc. (KKNA - Kyowa Kirin North America) and the Bodman family (TE), as well as the US Centers for Disease Control and Prevention (CDC) (cooperative agreement IP16-004), National Institutes of Health/NIAID (U01A1144673), and the Open Philanthropy/Good Ventures Foundation (GV673600847) (MAS).

## Supplementary Figures

**Figure S1.**
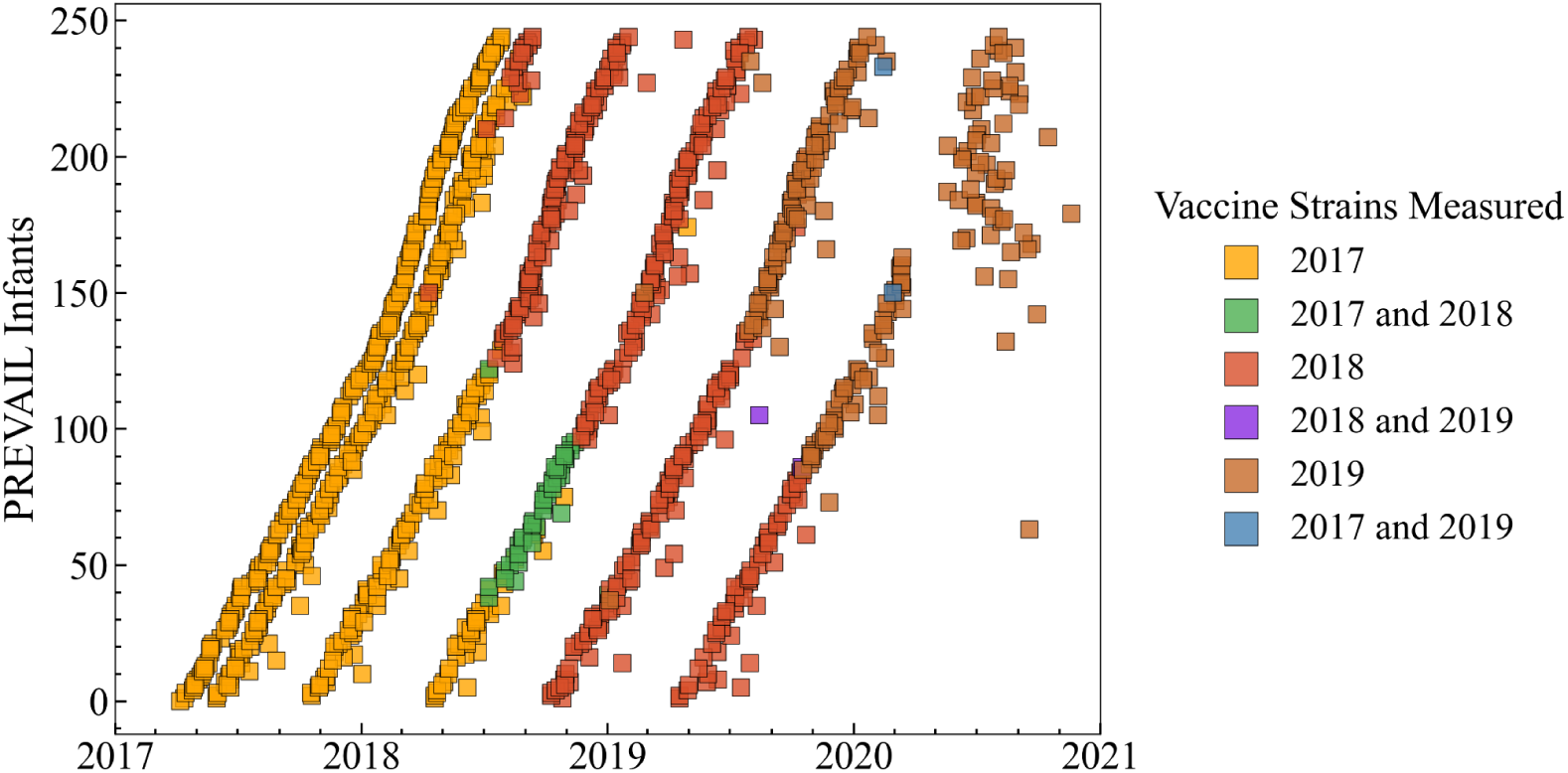
HAI virus panel for each infant and time point. The 2017 vaccine strains included H1N1 A/Michigan/45/2015, H3N2 A/Hong Kong/4801/2014, B Victoria B/Brisbane/60/2008, and B Yamagata B/Phuket/3073/2013. The 2018 vaccine strains included H1N1 A/Michigan/45/2015, H3N2 A/Singapore/INFIMH-16-0019/2016, B Victoria B/Colorado/6/2017, and B Yamagata B/Phuket/3073/2013. The 2019 vaccine strains included H1N1 A/Brisbane/2/2018, H3N2 A/Kansas/14/2017, B Victoria B/Colorado/6/2017, B Yamagata B/Phuket/3073/2013. Some sera were measured against vaccine strains from two years (green, purple, teal).

**Figure S2.**
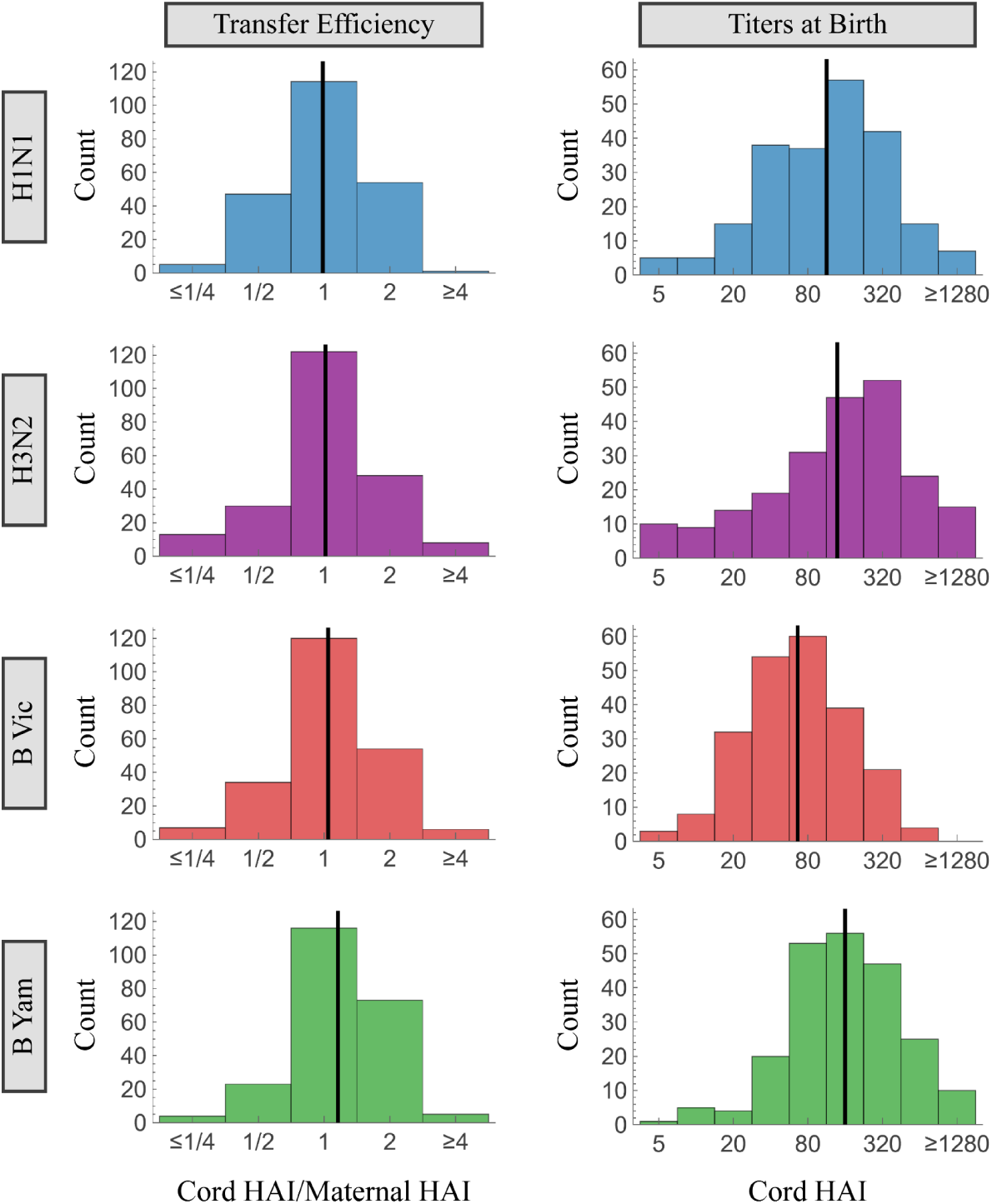
Antibody transfer efficiency and the distribution of cord blood HAI titers. *Left*: Maternal-to-infant antibody transfer efficiency defined as the ratio of maternal HAI titers to cord blood HAI titers. *Right*: Distribution of infant cord blood HAI titers. In each plot, the vertical line denotes the geometric mean of the distribution.

**Figure S3.**
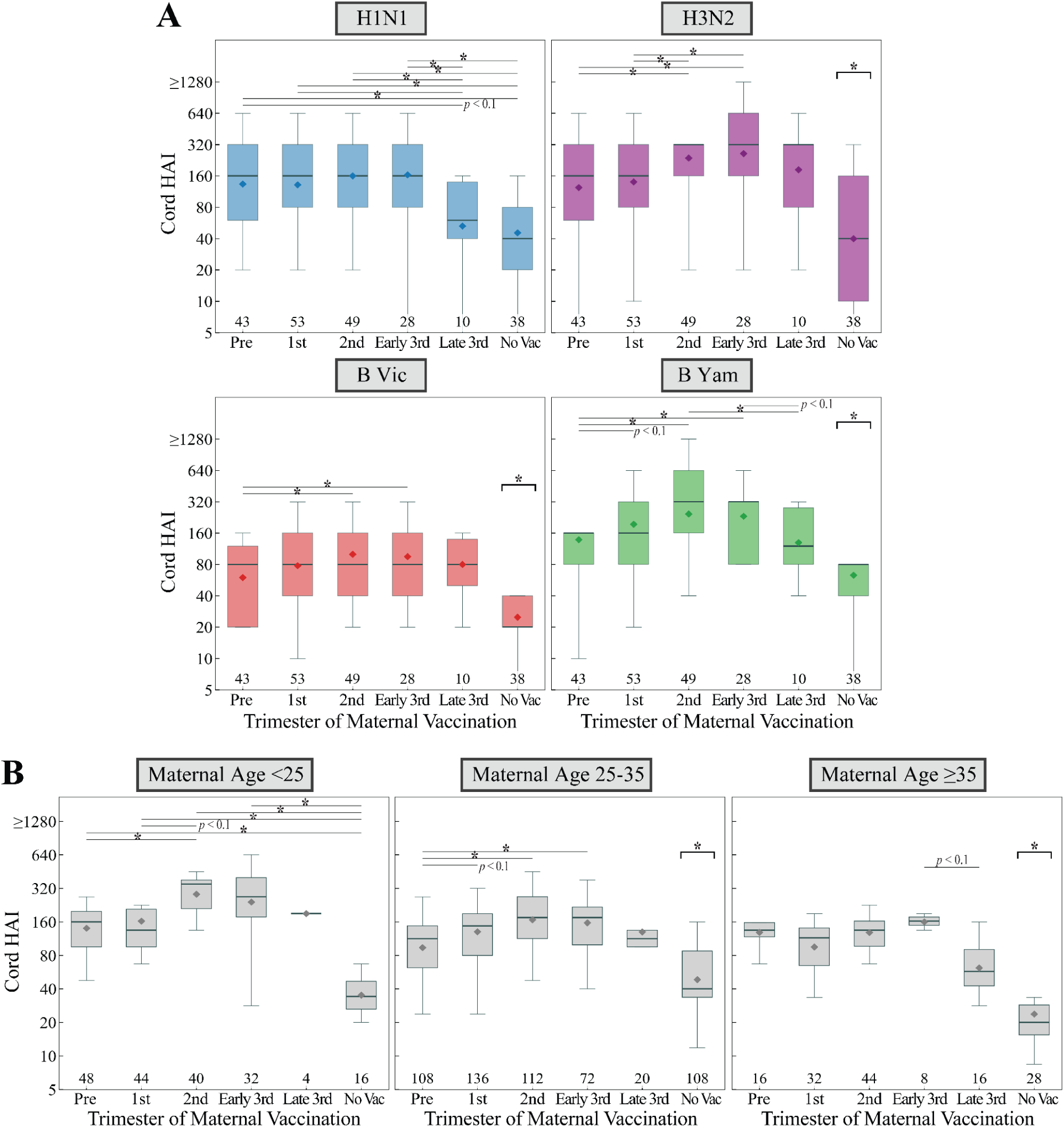
Infant cord blood HAI titers based on timing of maternal vaccination. (A) Infant cord blood HAI titers separated by individual vaccine strain (H1N1, H3N2, B Victoria, and B Yamagata). The values below each group indicate the number of mothers. (B) Combined cord blood HAI titers across all strains, stratified by maternal age groups (<25, 25–35, and ≥35 years). The number of measurements=(# of mothers)×(# of vaccine strains) is shown below each group. For all panels, box plots show the interquartile range, horizontal lines the medians, and black diamonds the GMTs. Whiskers extend to 1.5 times the interquartile range. Bracket indicates significance against all other groups. *: *p* ≤ 0.05 (Welch’s t-test on the log-transformed values).

**Figure S4.**
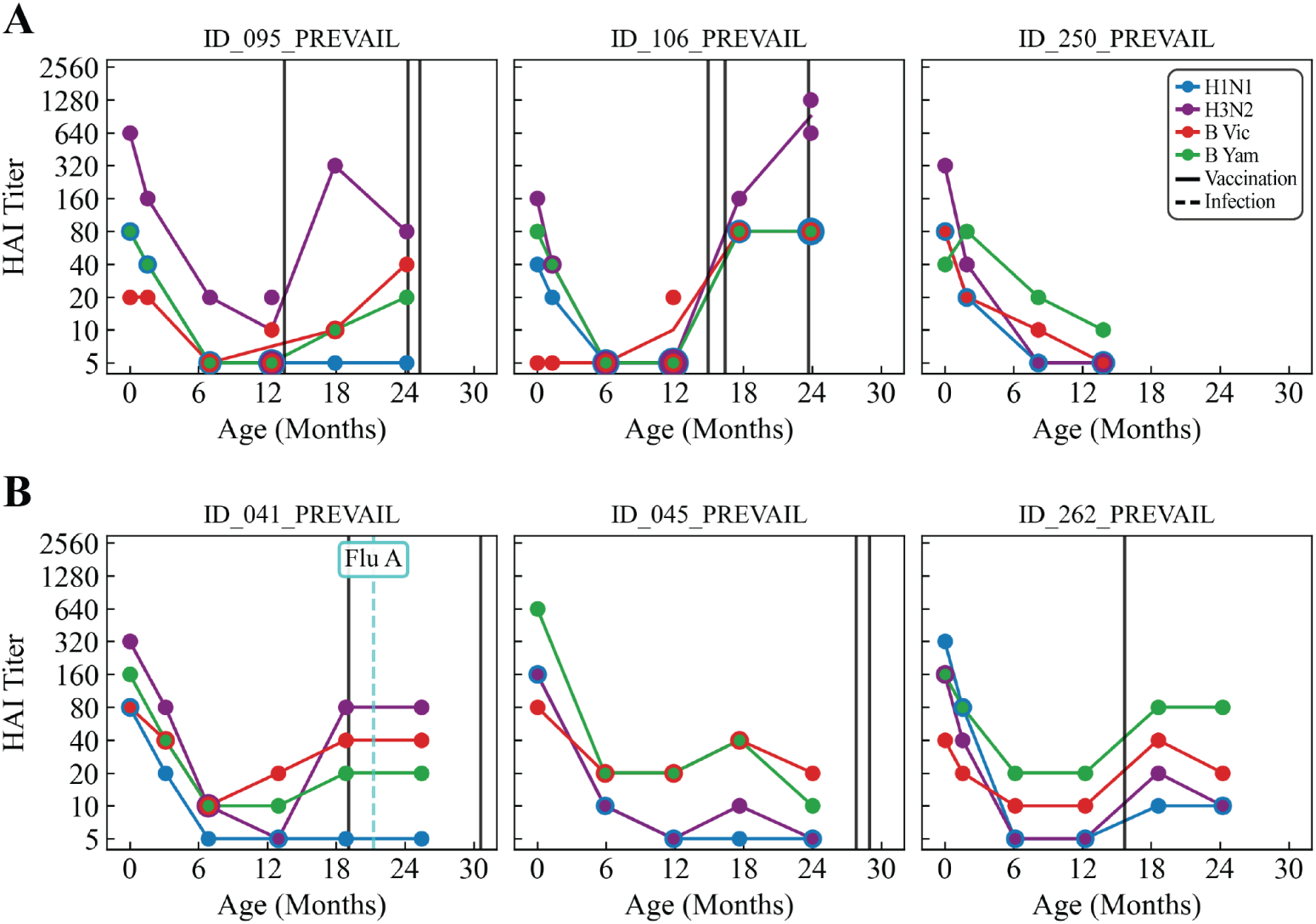
Representative HAI trajectories of children who were not vaccinated during their first year of life. (A) Children whose B Victoria or B Yamagata titers hit the limit of detection (HAI of 5) at 12 months. (B) Children whose influenza B titers remained above the limit of detection at 12 months before vaccination. In each longitudinal trajectory, black vertical lines represent vaccinations and dashed colored lines represent infections (*e.g.*, ID_041_PREVAIL had an influenza A infection with unknown subtype).

**Figure S5.**
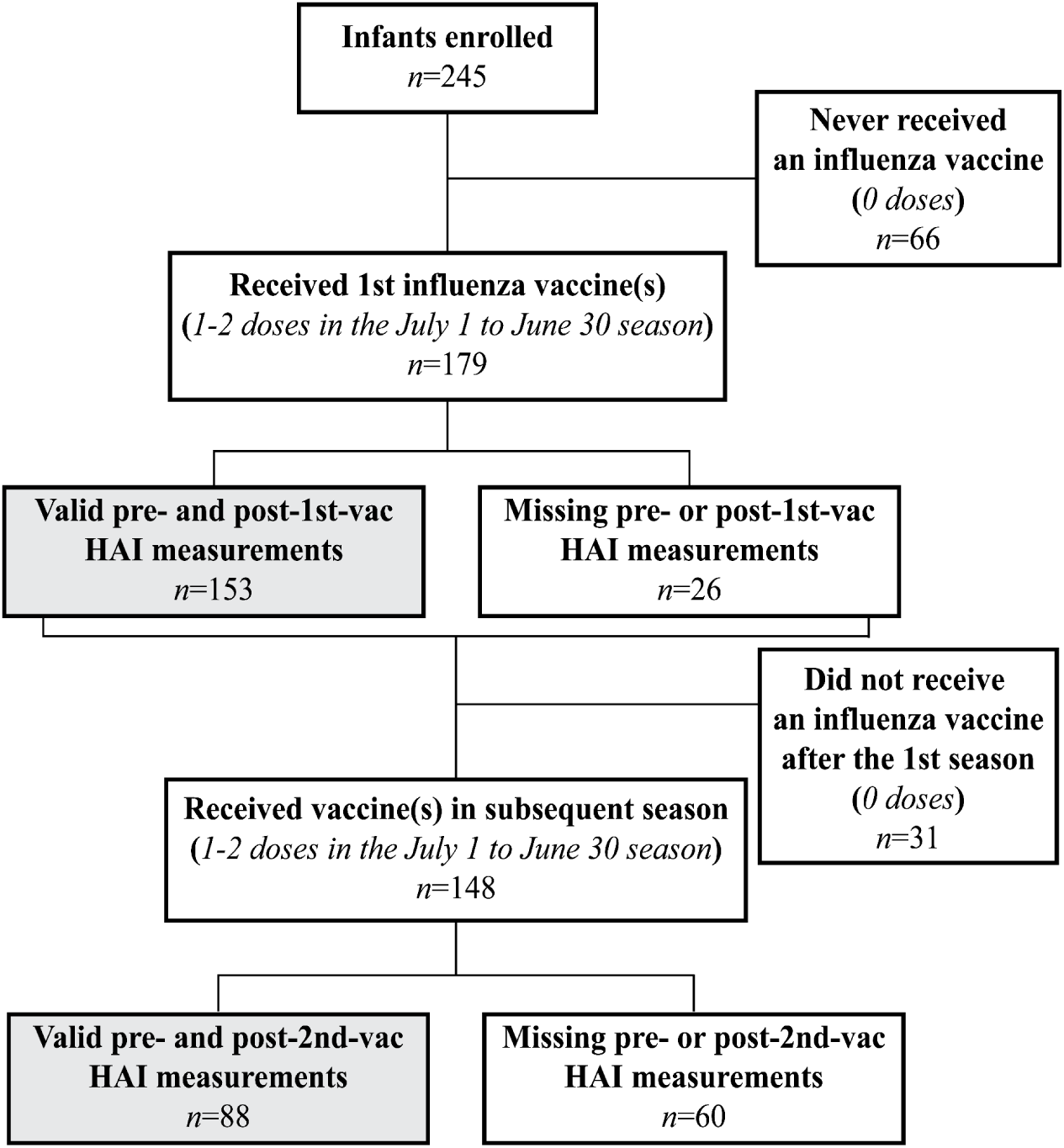
Flow chart for 1st and 2nd vaccination seasons. Participant retention from initial infant enrollment through the 2nd vaccination season. Gray boxes highlight the final sample sizes of infants with valid pre- and post-vaccination HAI measurements analyzed for the 1st and 2nd vaccination seasons.

**Figure S6.**
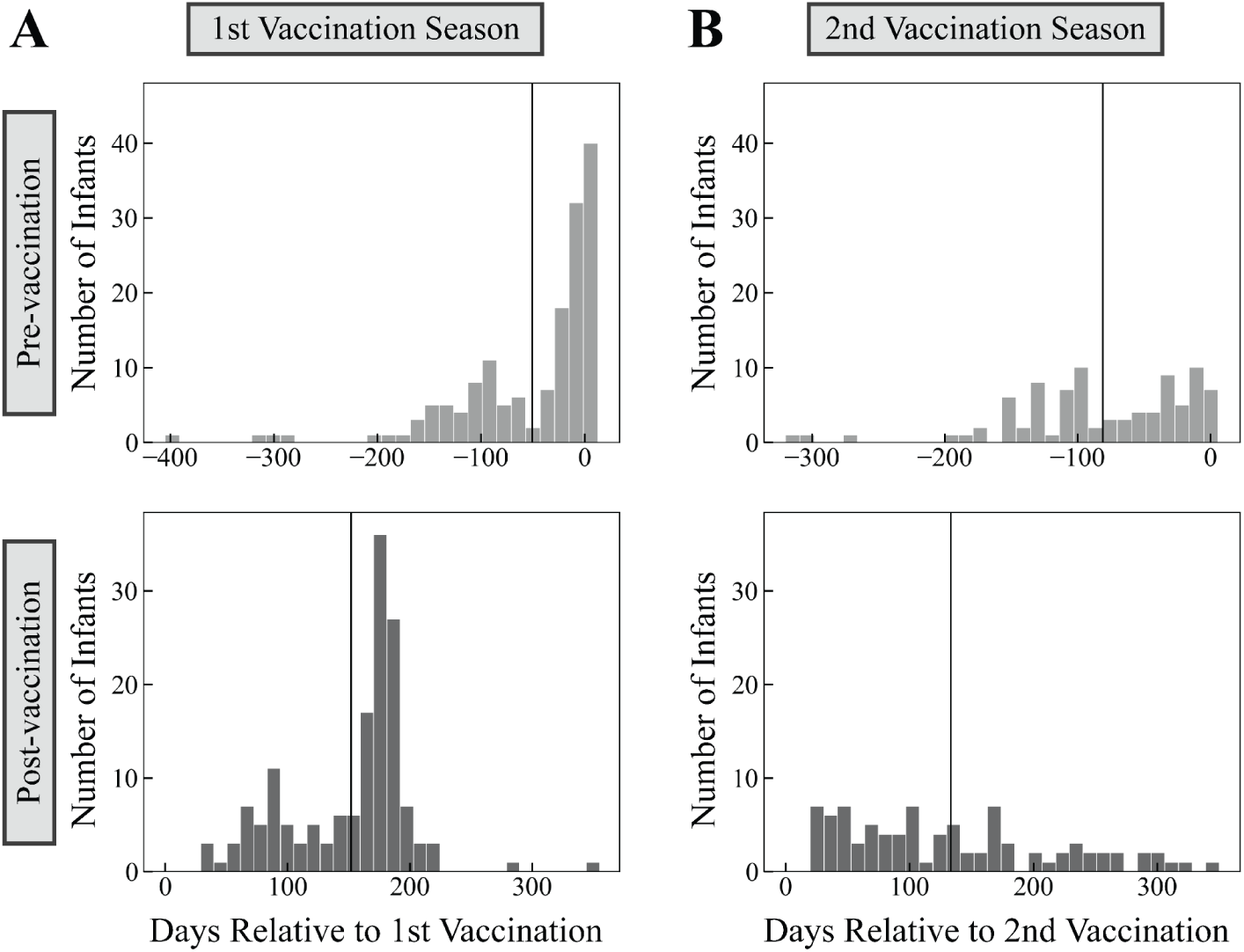
Number of days between pre-/post-vaccination serum collection and the date of vaccination. (A) To calculate fold-change for 1st vaccination season: Pre-vaccination was defined as the latest measurement up to 14 days post-1st-vaccination. Post-vaccination was defined as the earliest measurement between 28 days post-1st-vaccination and 7 days post-2nd-vaccination. Sera from 15-27 days were used to assess dynamics but not fold-change. (B) 2nd vaccination season: Pre-vaccination was defined as the latest measurement between 28 days post-1st-vaccination and 7 days post-2nd-vaccination. Post-vaccination was defined as the earliest measurement taken at least 14 days post-2nd-vaccination. Sera from 8-13 days were used to assess dynamics but not fold-change. Vertical lines indicate the mean collection day for each vaccination.

**Figure S7.**
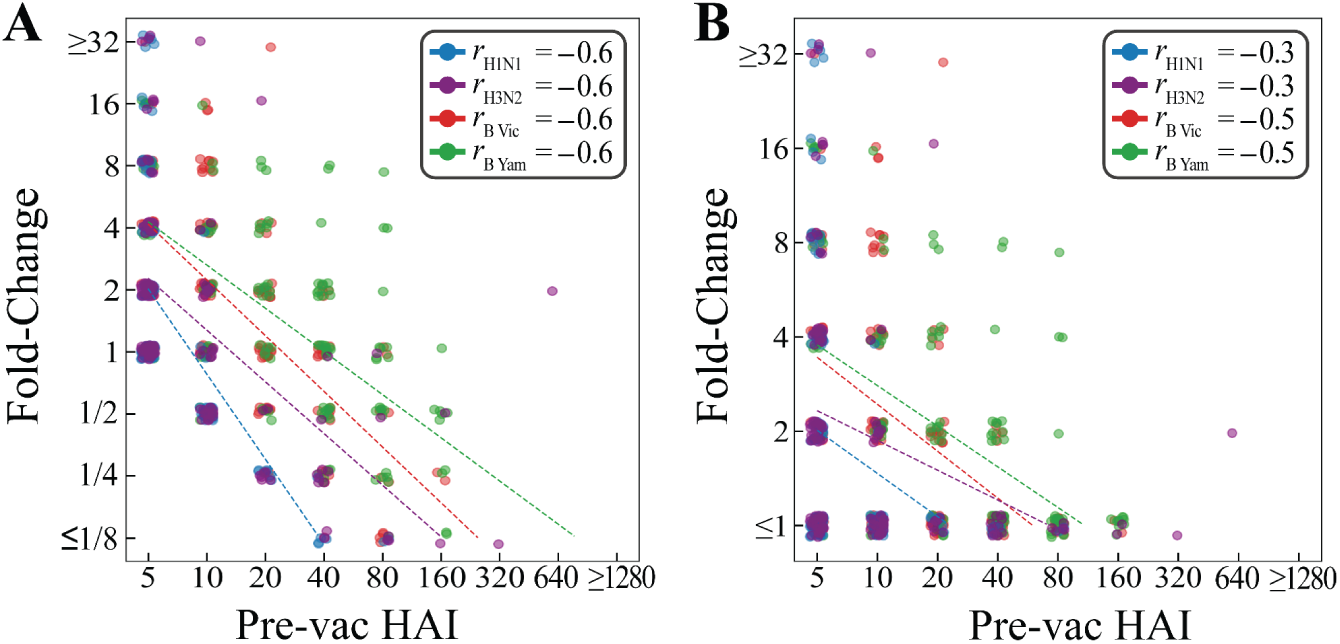
Antibody ceiling effect from the 1st infant vaccination. Pre-vaccination HAI titers vs fold-change shown when (A) HAI titers are rounded to the nearest standard titer value (5, 10, 20…) rather than the continuous range of imputed values. (B) In addition to rounding to the nearest 2-fold titer, fold-change was also clipped to be ≥1 to estimate the effects in older children where maternal antibody decay is no longer possible. In each case, linear regression was performed on each vaccine strain to quantify the antibody ceiling effect.

**Figure S8.**
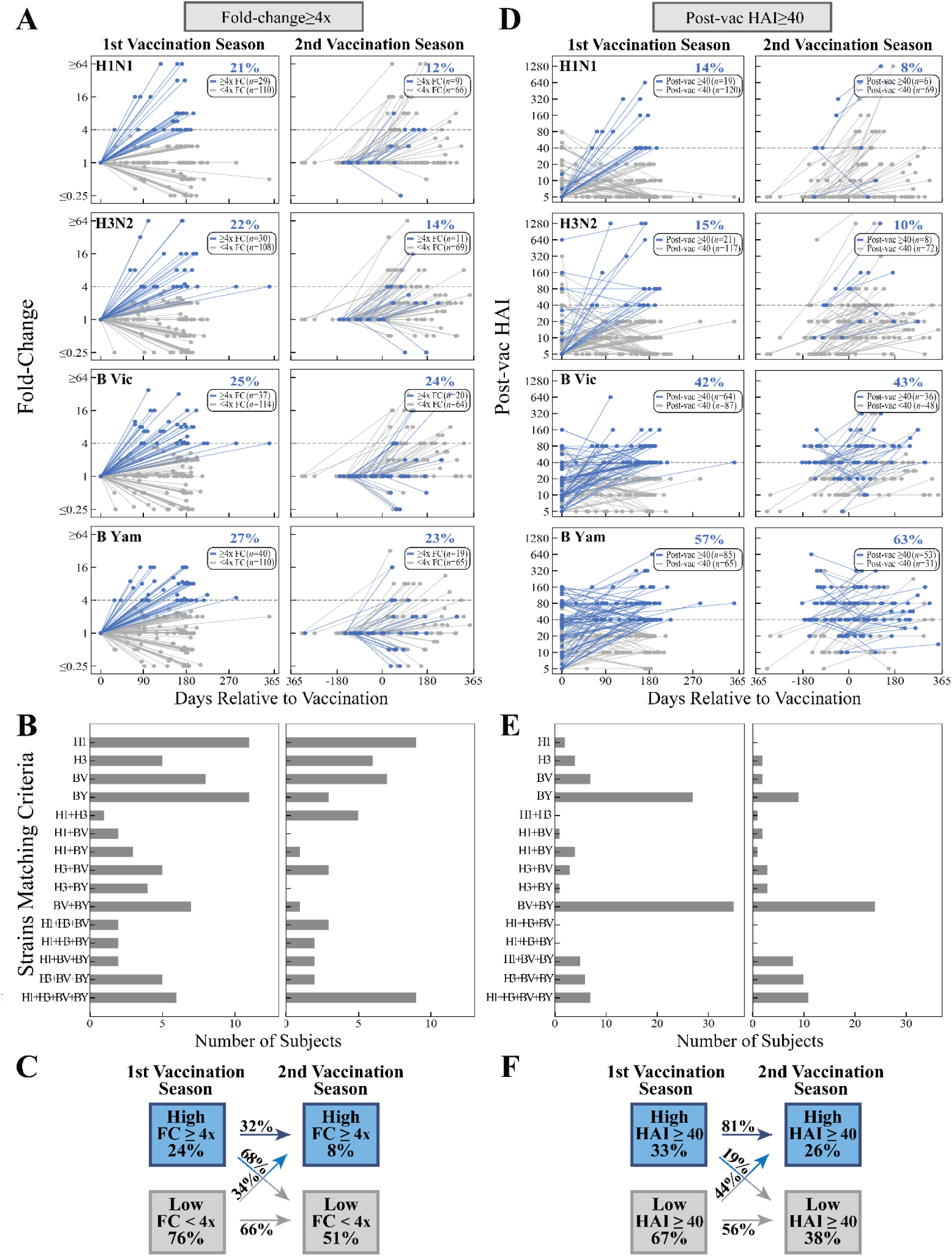
1st and 2nd vaccine responses with fold-change≥4x or post-vaccination HAI titer≥40. High responders (blue) are defined as having (A-C) fold-change≥4x or (D-F) post-vaccination HAI titer≥40. (A, D) For every vaccine strain, percent of HAI titer trajectories matching each criterion. (B, E) For each infant, combination of vaccine strains matching each criterion (*e.g.*, H1=only H1N1 satisfies criterion, H3+BY=only H3N2 and B Yamagata satisfy criterion). (C, F) Proportion of strain-specific responses matching the criteria (box), and the percentages of these responses that remained the same or changed between 1st and 2nd vaccination seasons (arrows).

**Figure S9.**
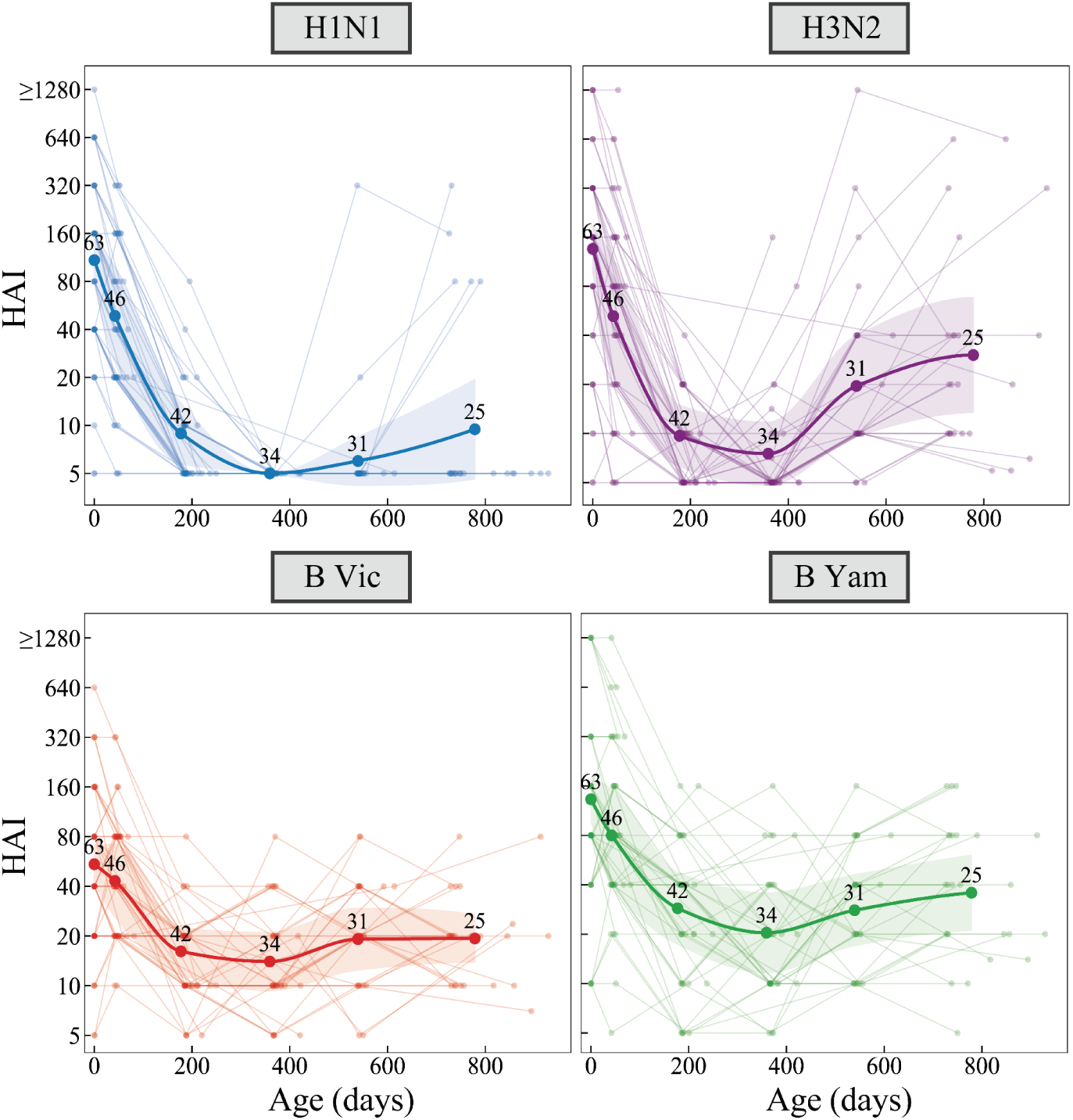
Longitudinal HAI titer trajectories in unexposed children. Trajectories of HAI titers against four vaccine strains (H1N1, H3N2, B Victoria, and B Yamagata) in children with no prior vaccination or infection. Thin lines represent individual child trajectories, while thick lines denote the overall trend. Data were binned into time points containing ≥10 children and connected using splines. The numerical values above the trendlines specify the exact sample size for each bin (**Methods**).

**Figure S10.**
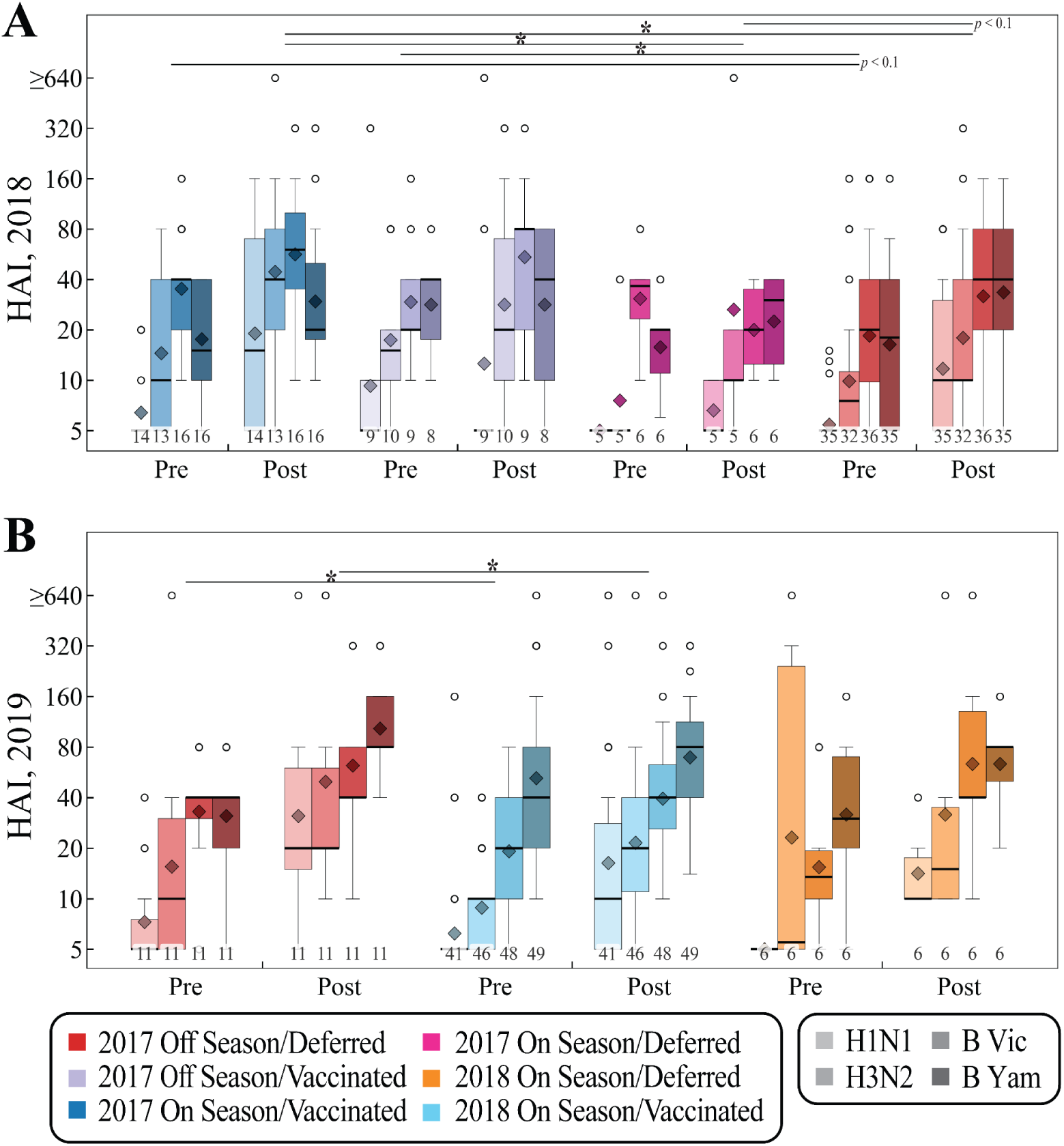
Vaccine-elicited antibody titers for the infants in 2018 and 2019 influenza season. Pre-and post-vaccination titers for the (A) 2018 and (B) 2019 season (analogous to **Fig 3C,F** but showing the four vaccine strains H1N1, H3N2, B Victoria, and B Yamagata from left-to-right). All box plots show the interquartile range, horizontal lines the medians, and dark diamonds the geometric mean titers. Whiskers extend to 1.5 times the interquartile range. The number of infants measured for each subtype is shown below each box plot. *: p ≤ 0.05 (Welch’s t-test on the log-transformed values, Stouffer combination across strains).

**Figure S11.**
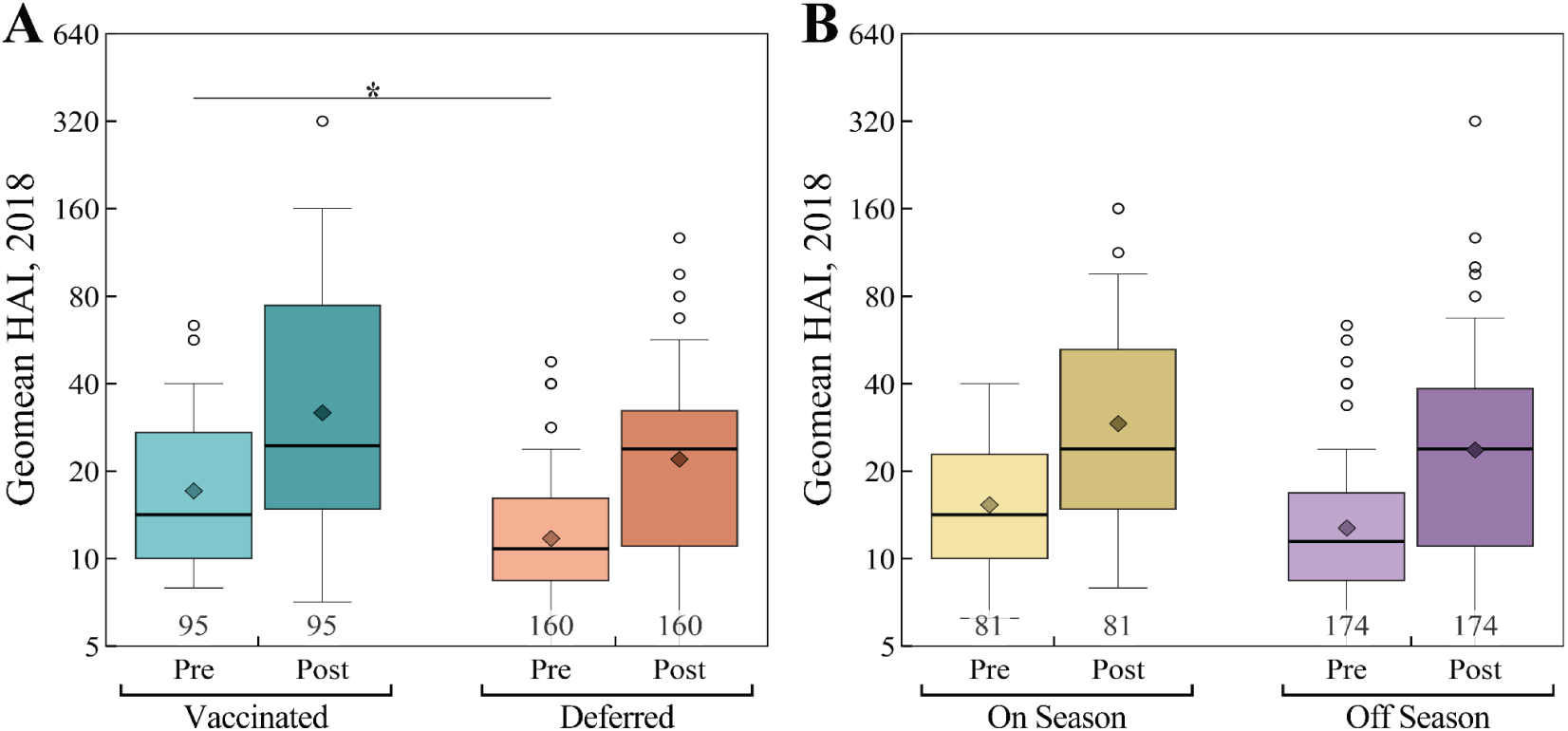
HAI titer distributions in 2018 for different groups of infants. (A) All vaccinated infants (on- or off-season, **Fig 3B** blue/violet) vs all deferred infants (on- or off-season, **Fig 3B** red/pink). (B) All on-season infants (vaccinated or deferred, **Fig 3B** blue/pink) vs all off-season infants (on- or off-season, **Fig 3B** violet/red). *: *p* ≤ 0.05 (Welch’s t-test on the log-transformed values)

**Figure S12.**
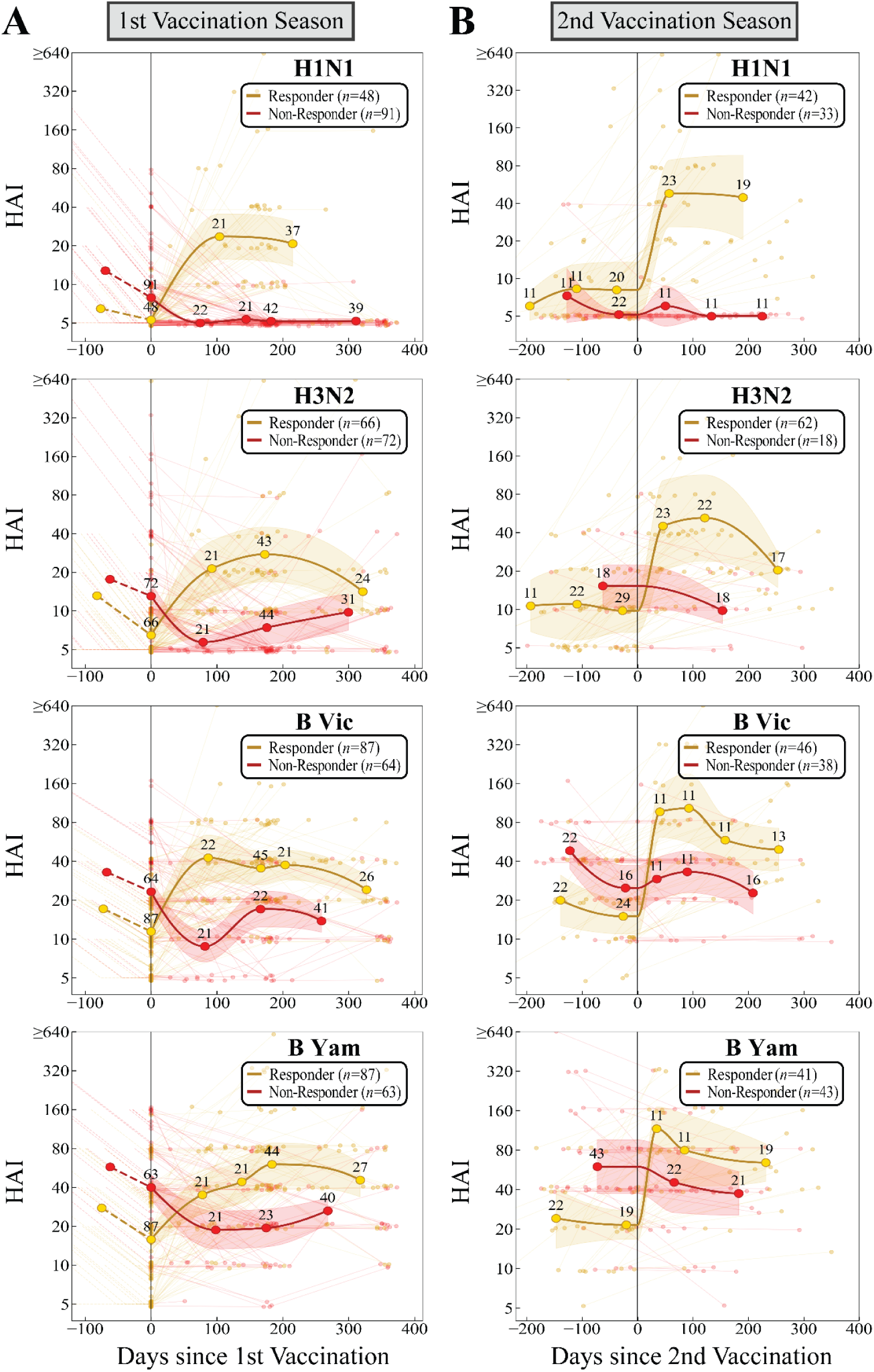
High-resolution dynamics of 1st and 2nd vaccine response. HAI trajectories for responders (titers increase post-vaccination) and non-responders (all others) during the (A) 1st or (B) 2nd vaccination seasons. Responder/non-responder phenotypes are assessed independently for each vaccine strain and vaccination. Time points were binned to include a minimum of 10 children (≥10) and connected using splines. The exact sample size is indicated above each point (**Methods**). Rows from top-to-bottom represent H1N1, H3N2, B Victoria, and B Yamagata.

**Figure S13.**
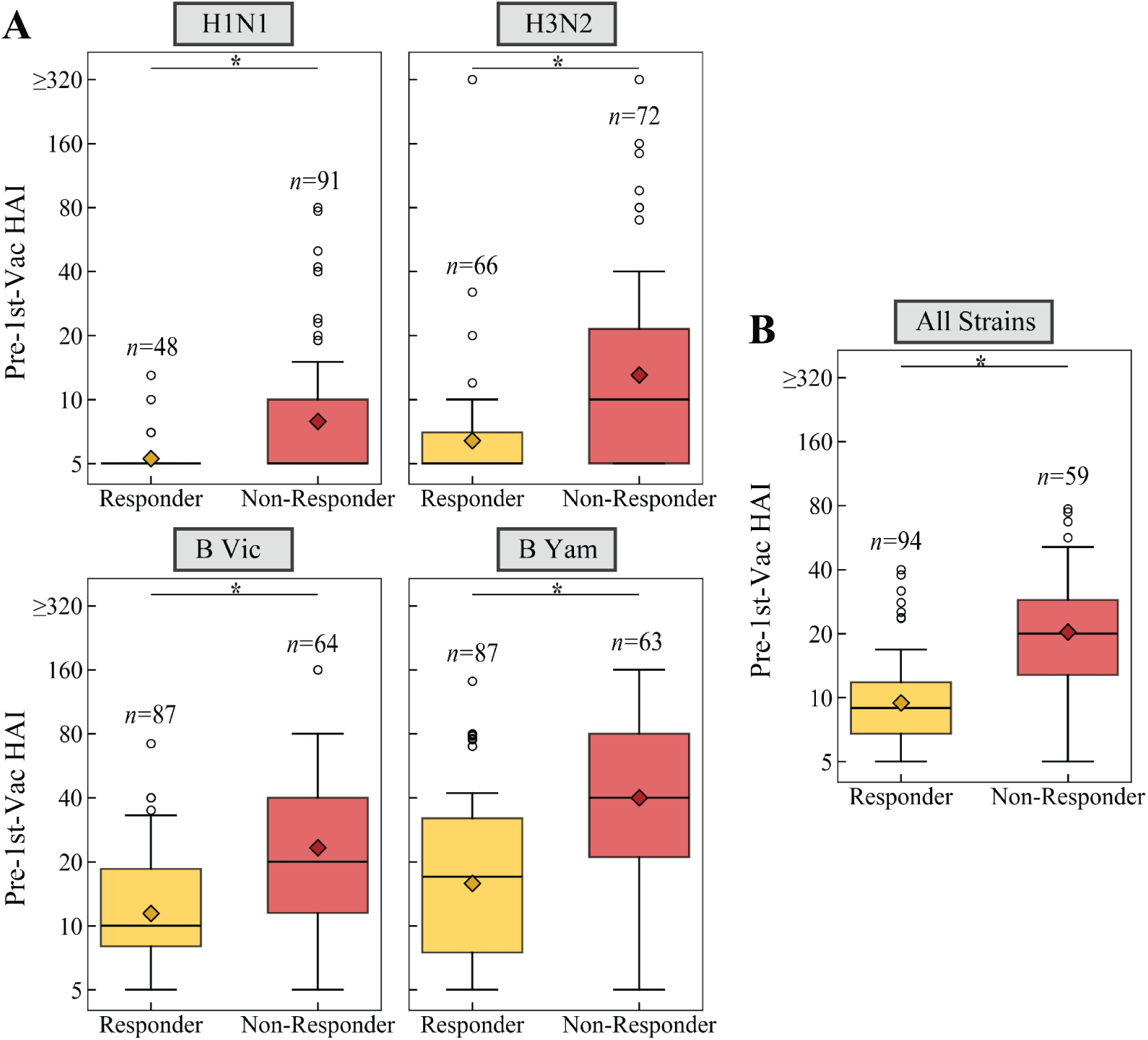
Features of 1st vaccine responder and non-responder phenotypes. Pre-vaccination HAI titers against (A) each vaccine strain or (B) the geomean titer against all vaccine strains. Responders (titers increase post-vaccination) and non-responders (all others) are defined independently for each vaccine strain. *: p ≤ 0.05 (Welch’s t-test on the log-transformed values)

**Figure S14.**
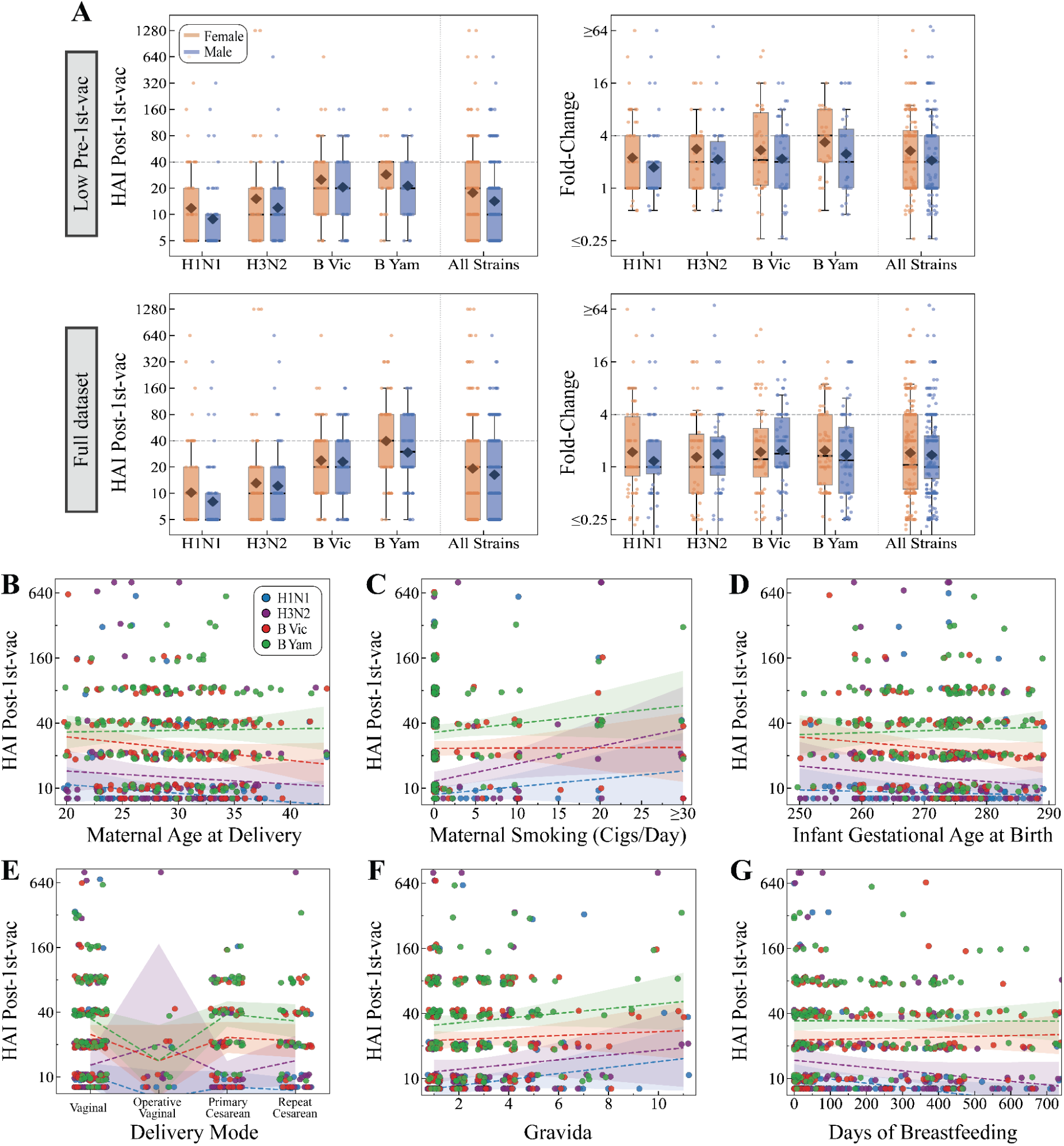
Dependence of post-vaccination HAI titers on maternal and infant features. (A) Comparison of post-1st-vac HAI titers (left) and fold-change (right) between female and male infants. Data are presented for infants with low pre-vaccination titers (top row) and the full dataset (bottom row) across specific influenza strains (H1N1, H3N2, B Victoria, B Yamagata) and all strains combined. Each infant’s post-1st-vaccination HAI titer for each vaccine strain is compared against (B) maternal age at delivery [years], (C) maternal smoking pre-pregnancy [cigarettes/day], (D) infant gestational age [days], (E) delivery mode, (F) gravida, and (G) days of breastfeeding [either exclusive or partial breastfeeding]. Dashed lines indicate strain-specific trend lines, and shaded bands denote 95% confidence intervals.

**Figure S15.**
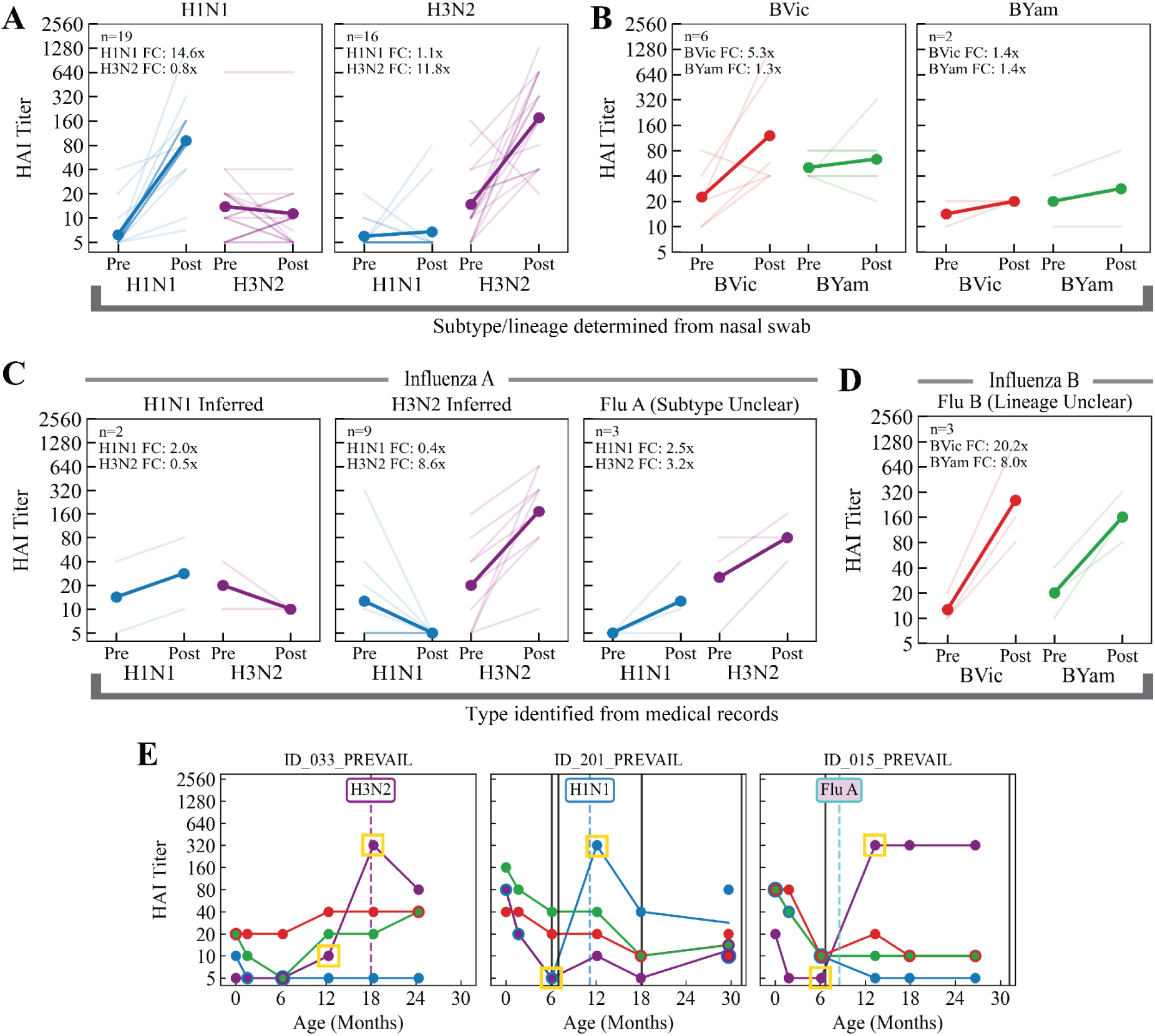
Subclassification of influenza virus infections. (A) Comparison of H1N1 and H3N2 HAI titers between pre- and post-infection timepoints for infections subtyped through nasal swabs. (B) Comparison of B Yamagata and B Victoria titers for infections with known lineage through nasal swabs. (C) Classification of influenza A infections that were typed (but not subtyped) through medical chart data. Cases were categorized as “H1N1 Inferred” when titers increased for H1N1 but not H3N2, with an analogous definition for “H3N2 Inferred.” All other cases were denoted by “Flu A (Subtype Unclear).” (D) Influenza B infections that were typed (with no lineage specified) could not be further classified since titers to both B lineages increased post-infection. (E) Representative HAI trajectories from individuals with an H3N2 or H1N1 infection subtyped through nasal swabs as well as an influenza A infection whose subtype could be determined serologically as an H3N2 infection (and thus the “Flu A” box is filled in using purple H3N2 coloring). Yellow boxes indicate the pre- and post-infection measurements used in the analyses for these three children.

**Figure S16.**
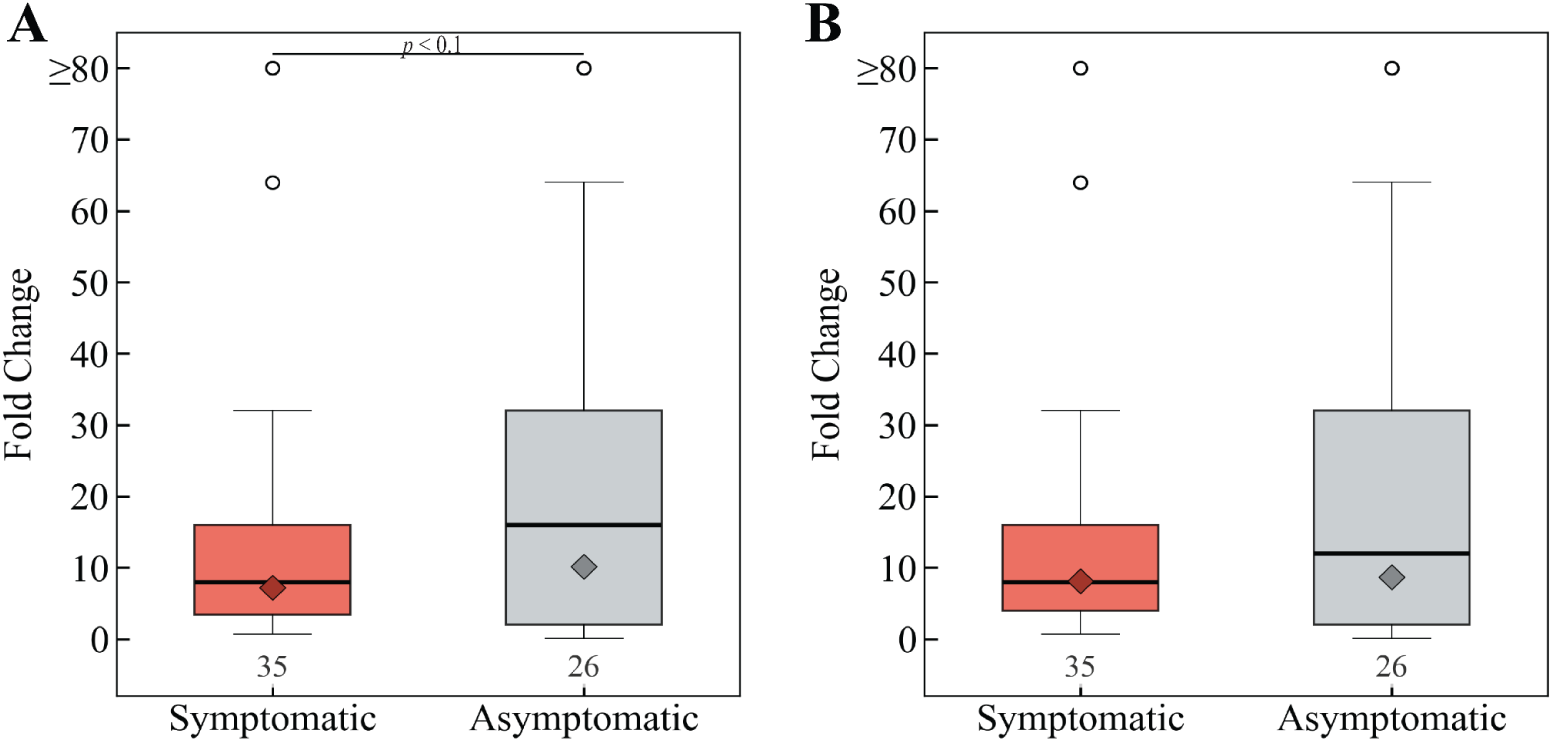
Comparison of antibody responses between symptomatic and asymptomatic infections. (A) Children were classified as *Symptomatic* if any of the influenza-related symptoms (fever, cough, vomiting, diarrhea, dehydration, or altered mental status) were recorded in *medical charts* within ±10 days of the positive swab. (B) Children were classified as *Symptomatic* if the positive nasal swab date fell within an acute respiratory illness episode recorded via weekly *maternal text reports*. P-value: Welch’s t-test on the log-transformed values.

